# Deaths averted by COVID-19 vaccination in select Latin American and Caribbean Countries: a modelling study

**DOI:** 10.1101/2024.04.12.24305739

**Authors:** Alexandra Savinkina, Daniel M. Weinberger, Cristiana M. Toscano, Lucia H. De Oliveira

**Affiliations:** Department of Epidemiology of Microbial Diseases and Public Health Modeling Unit, Yale School of Public Health, Yale University; New Haven, CT USA; Federal University of Goias, Institute of Tropical Pathology and Public Health, Goiania-GO, Brazil; Comprehensive Immunization Program, Pan-American Health Organization (PAHO), Washington – DC, USA

**Author notes:** **Corresponding Author:** Alexandra Savinkina 60 College Street New Haven, CT 06520-8034. The author is a staff member of the Pan American Health Organization. The author alone is responsible for the view expressed in this publication, and they do not necessarily represent the decisions or policies of the Pan American Health Organization.

**Keywords:** Covid-19, Latin America, Caribbean, Vaccination, Mathematical Modeling

## Abstract

**Background:** The COVID-19 pandemic has had a significant impact on global health, with millions of lives lost worldwide. Vaccination has emerged as a crucial strategy in mitigating the impact of the disease. This study aims to estimate the number of deaths averted through vaccination in LAC during the first year and a half of vaccination rollout (January 2021 - May 2022).

**Methods:** Publicly available data on COVID-19 deaths and vaccination rates were used to estimate the total number of deaths averted via vaccination in LAC. Using estimates for number of deaths, number of vaccinated, and vaccine effectiveness, a counterfactual estimated number of deaths observed without vaccination was calculated. Vaccine effectiveness estimates were obtained from published studies. The analysis focused on 17 countries in LAC and considered adults aged 18 years and above.

**Findings:** After accounting for underreporting, the analysis estimated that over 1.49 million deaths were caused by COVID-19 in the selected countries during the study period. Without vaccination, the model estimated that between 2.10 and 4.11 million COVID-19 deaths would have occurred. Consequently, vaccination efforts resulted in approximately 610,000 to 2.61 million deaths averted.

**Interpretation:** This study represents the first large-scale, multi-center estimate of population-level vaccine impact on COVID-19 mortality in LAC. The findings underscore the substantial impact of timely and widespread vaccination in averting COVID-19 deaths. These results provide crucial support for vaccination programs aimed at combating epidemic infectious diseases in the region and future pandemics.

**Funding:** This study was funded by the Pan-American Health Organization (PAHO).

## Introduction

The World Health Organization (WHO) declared a public health emergency of international concern (PHEIC) on January 30, 2020 in response to the identification of a novel coronavirus (SARS-CoV-2) in China. In March 2020, as lab-confirmed SARS-CoV-2 cases exponentially grew in China, and newly identified transmission was reported in other countries, WHO declared the outbreak a global pandemic and called on countries to rapidly respond with control and mitigation plans to slow the spread of the virus. In the ensuing months, countries worldwide have faced challenges to keep a responsive pace with the spread of the virus, which has led to substantial health and socioeconomic losses and significant mortality related to COVID-19 worldwide.^1^ To date, a total of 7 million deaths due to COVID-19 have been reported globally since January 1^st^ 2020.

Countries in the Americas have been among the hardest hit by the pandemic. By early 2023, approximately 43% of all reported COVID-19 deaths in the world had occurred in the region, reaching 2.89 million deaths as of January 1^st^, 2023. To date, South America as a sub-region experienced 1.35 million COVID-19 deaths during the pandemic.^2^ Although most COVID-19 deaths occur among older adults and individuals living with comorbidities, deaths have occurred in all ages including young children, even though younger ages appear to be less susceptible to severe disease.

The first vaccines to prevent COVID-19 became available for use in early 2021. As of July 2021, eight COVID-19 vaccines had received Emergency Use Listing (EUL) by the WHO pre-qualification process,^3^ upon meeting predefined criteria for safety and efficacy. By May 2023, this number had reached 15,^4^ and many more are under assessment for pre-qualification.^5^

The rapid deployment of vaccines has been proven critical to halt the pandemic’s toll in the region. Many countries in the region accessed vaccines via PAHO’s Revolving Fund, in active collaboration with the COVAX Facility. In Latin American and Caribbean (LAC) countries, 82% of the population has received at least one dose of COVID-19 vaccine to date, and 1.1 billion doses of vaccine have been administered.^6^ Despite the initial availability of COVID-19 vaccines across the region, wide inter- and intra-country variation in access and availability to vaccines were observed in the region.^6^

Selected studies have recently used models to estimate the impact of COVID-19 vaccination in averting COVID-19 deaths in selected countries and periods.^7,8^ To date, the burden of COVID-19 deaths prevented by vaccination, a key measure of the impact of the vaccination program, has not been established for the Latin American region. This study aims to estimate the numbers of COVID-19 deaths averted due to vaccination in selected countries of the LAC region during the COVID-19 pandemic.

## Methods

### Study design, setting and period

We used existing data on reported deaths due to COVID-19 and COVID-19 vaccination coverage over time to estimate deaths averted via vaccination for select countries in Latin America and the Caribbean. This modelling study used data from 17 countries in the LAC region during the period ranging from vaccine introduction in each country (early 2021) to May 2022 (Table 1). The 17 countries were selected for inclusion because they had available COVID-19 vaccination coverage data (Figure 1). This included: Argentina, Brazil, Chile, Colombia, Paraguay, Uruguay, Jamaica, Peru, Belize, Bolivia, Costa Rica, Ecuador, El Salvador, Guatemala, Honduras, Mexico, and Venezuela. All other countries in the Latin American and Caribbean regions were excluded due to missing data on COVID-19 vaccination coverage and/or COVID-19 deaths. Our analysis considered only adults over 18, and the model was stratified by age (18-59 years old, 60 years and older).

**Figure 1.**
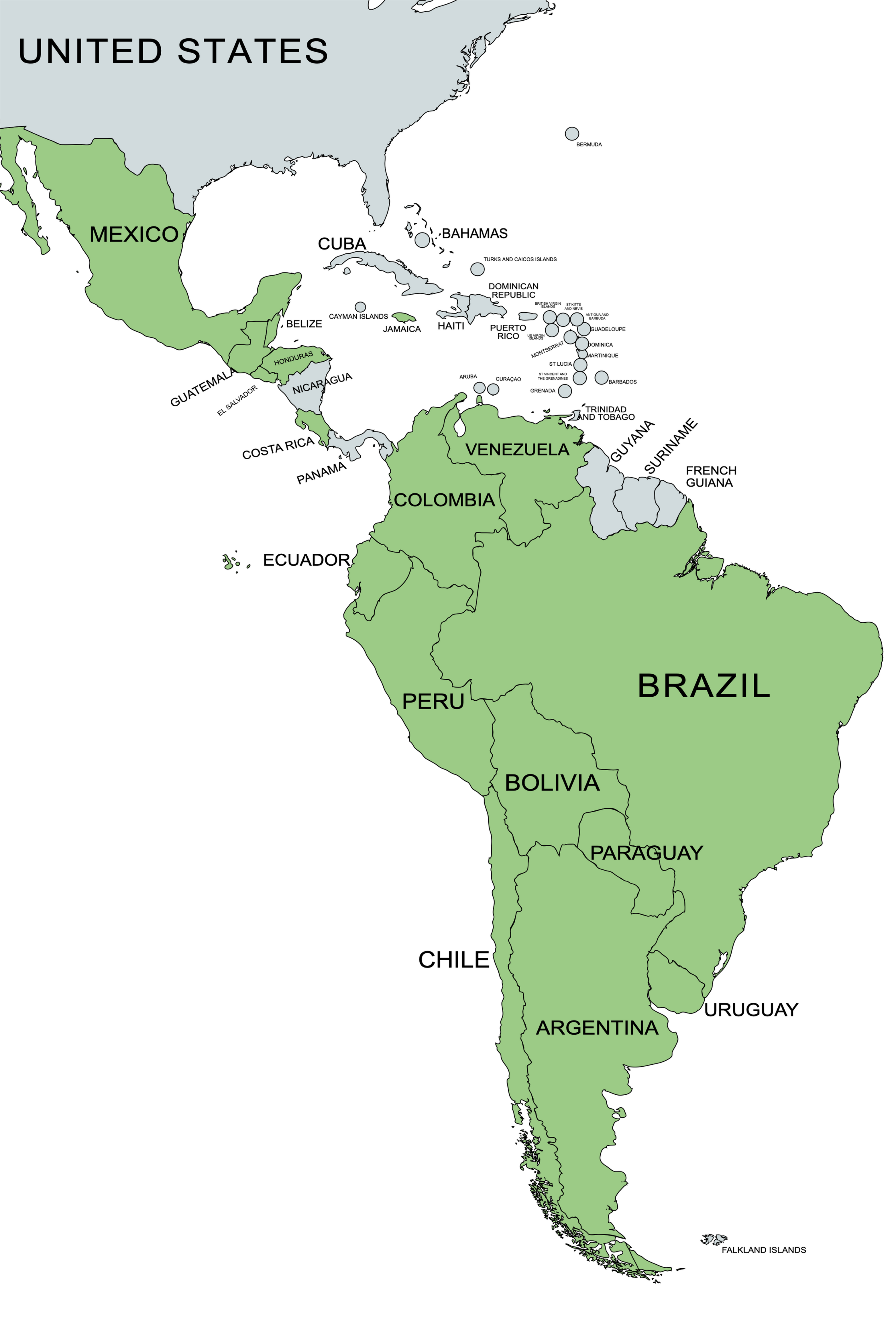
Map showing 17 countries in the Latin American and Caribbean regions which were included in the modeling analysis. Included countries are shown in grey and excluded countries in white. Included countries are: Argentina, Brazil, Chile, Colombia, Paraguay, Uruguay, Jamaica, Peru, Belize, Bolivia, Costa Rica, Ecuador, El Salvador, Guatemala, Honduras, Mexico, and Venezuela.

**Table 1.** Country-specific data on vaccination, including start date of vaccination, percent coverage by age-groups considered in the analysis, and vaccines used in period.

| Country | Start date of vaccination | % over 60 yo vaccinated by May 2022 | % 18-59 yo vaccinated by May 2022 | Vaccines in use through May 2022<sup>1</sup> | 1. Source: Our World in Data a COVID-19 vaccine database<sup>1 3</sup> |
| --- | --- | --- | --- | --- | --- |
| Argentina | 2020-12 | 95 | 90.8 | CanSino, Moderna, Oxford/AstraZeneca, Pfizer/BioNTech, Sinopharm/Beijing, Sputnik V |  |
| Brazil | 2021-01 | 98.4 | 85.9 | Johnson&Johnson, Pfizer/BioNTech, Oxford/AstraZeneca, Sinovac |  |
| Chile | 2020-12 | 93.9 | 96.9 | CanSino, Moderna, Oxford/AstraZeneca, Pfizer/BioNTech, Sinovac |  |
| Colombia | 2021-03 | 81.1 | 48.3 | Johnson&Johnson, Moderna, Oxford/AstraZeneca, Pfizer/BioNTech, Sinovac |  |
| Paraguay | 2021-02 | 76 | 59.9 | Covaxin, Moderna, Oxford/AstraZeneca, Pfizer/BioNTech, Sinopharm/Beijing, Sinovac, Sputnik V |  |
| Uruguay | 2021-03 | 100 | 79.8 | Sinovac, Pfizer/BioNTech, Oxford/AstraZeneca |  |
| Jamaica | 2021-03 | 35.2 | 26.4 | Johnson&Johnson, Moderna, Pfizer/BioNTech, Oxford/AstraZeneca |  |
| Peru | 2021-02 | 90.2 | 91.1 | Moderna, Oxford/AstraZeneca, Pfizer/BioNTech, Sinopharm/Beijing |  |
| Belize | 2021-02 | 74.7 | 67.8 | Johnson&Johnson, Oxford/AstraZeneca, Pfizer/BioNTech, Sinopharm/Beijing |  |
| Bolivia | 2021-02 | 70.9 | 62.6 | Johnson&Johnson, Oxford/AstraZeneca, Pfizer/BioNTech, Sinopharm/Beijing, Sputnik V |  |
| Costa Rica | 2021-02 | 85.1 | 90.4 | Oxford/AstraZeneca, Pfizer/BioNTech |  |
| Ecuador | 2021-02 | 90.5 | 87.1 | CanSino, Oxford/AstraZeneca, Pfizer/BioNTech, Sinovac |  |
| El Salvador | 2021-02 | 84.2 | 80.8 | Oxford/AstraZeneca, Pfizer/BioNTech, Sinopharm/Beijing, Sinovac |  |
| Guatemala | 2021-02 | 58.8 | 47.6 | Moderna, Oxford/AstraZeneca, Pfizer/BioNTech, Sputnik V |  |
| Honduras | 2021-02 | 66.4 | 34.6 | Johnson&Johnson, Moderna, Oxford/AstraZeneca, Pfizer/BioNTech, Sputnik V |  |
| Venezuela | 2021-02 | 42.2 | 58 | Abdala, Sinopharm/Beijing, Sinovac, Soberana02, Sputnik Light, Sputnik V |  |
| Mexico | 2020-12 | 92.7 | 80.1 | CanSino, Johnson&Johnson, Moderna, Oxford/AstraZeneca, Pfizer/BioNTech, Sinovac, Sputnik V |  |

### Data Sources

#### COVID-19 deaths

When possible, we used country-specific data for observed deaths over time from the COVerAGE-DB global demographic database of COVID-19 deaths.^9^ This dataset used data as reported by government entities, such as health ministries, and harmonized the data to standard metrics, measures, and age-bands using the penalized composite link model for ungrouping.^10^ The COVerAGE-DB database provided deaths by age group (18-59, 60+) as well as by time period (collapsed into monthly death counts). COVerAGE-DB was used for observed death counts in Argentina, Brazil, Chile, Colombia, Paraguay, Uruguay, Jamaica, and Peru. Mexico had COVID-19 mortality data available by age and time in COVerAGE-DB through October 2021, and this data was used for the analysis from December 2020 to October 2021. For countries that did not have COVID-19 death data reported in COVerAGE-DB either for the full time period of the analysis or for a part of it, we used non-age stratified COVID-19 death estimates over time from the World Health Organization Coronavirus (COVID-19) Dashboard.^1^ Data from the World Health Organization (WHO) was not age-stratified, therefore we imputed age stratified deaths using a linear regression model with population size by age category, country World Bank income level, and vaccination level in those over 60 as predictors. The linear regression model was validated using countries for which age-stratified death data were available. Additional methods can be found in the supplement. Countries for which death data were imputed from WHO reported numbers for the full time period of the analysis were Belize, Bolivia, Costa Rica, Ecuador, El Salvador, Guatemala, Honduras, and Venezuela. In addition, age-specific death counts were imputed for Mexico from November 2021 to May 2022. To account for the underreporting of COVID-19 deaths in the region, we used a multiplier considering available evidence from the literature, considering country-specific mortality underestimates ranging from 0 to 79% in studied countries (see Additional Methods).^11^

#### Vaccine coverage and effectiveness data

Data on vaccination by age over time by country and age group were obtained from the Pan-American Organization (PAHO)^12^ and from Our World in Data (OWID)^13^. Vaccine coverage considered partial and complete primary series, and first booster dose (not the second booster and beyond).

OWID data were complete for all countries of interest but were not age-stratified and included vaccination in those under 18, which was outside our study population. Data reported to PAHO were age-stratified, available as of September 2021, and less complete for some countries. Therefore, we used PAHO data to determine proportion of all vaccinations that were administered to each age group, and OWID to determine proportion of population within each age group that was vaccinated over time. Data for age-specific vaccination was only available for Mexico starting in February of 2022, we therefore assumed the same proportion of vaccines going to each age group in the months from December 2020 through January 2022 as in February 2022. As it is likely that elderly people were more prioritized for early vaccination, it is likely that this leads us to slightly underestimate^14–16^ the number of deaths averted in the older age group in the early part of 2021.

Multiple vaccine products were available and used by countries within PAHO over the course of vaccine administration (Table 1), and it is difficult to ascertain what proportion of all vaccinations were using specific products at each time point. Therefore, we considered a range of vaccine effectiveness (stratified by partial vs full vs booster vaccination) based on the vaccine effectiveness estimated for the vaccine products in the literature, with an effort to use studies conducted in Latin American countries.^17–26^ Additional boosters beyond the first were not considered in the analysis. Estimates of vaccine effectiveness can be seen in Table 2.

**Table 2.**
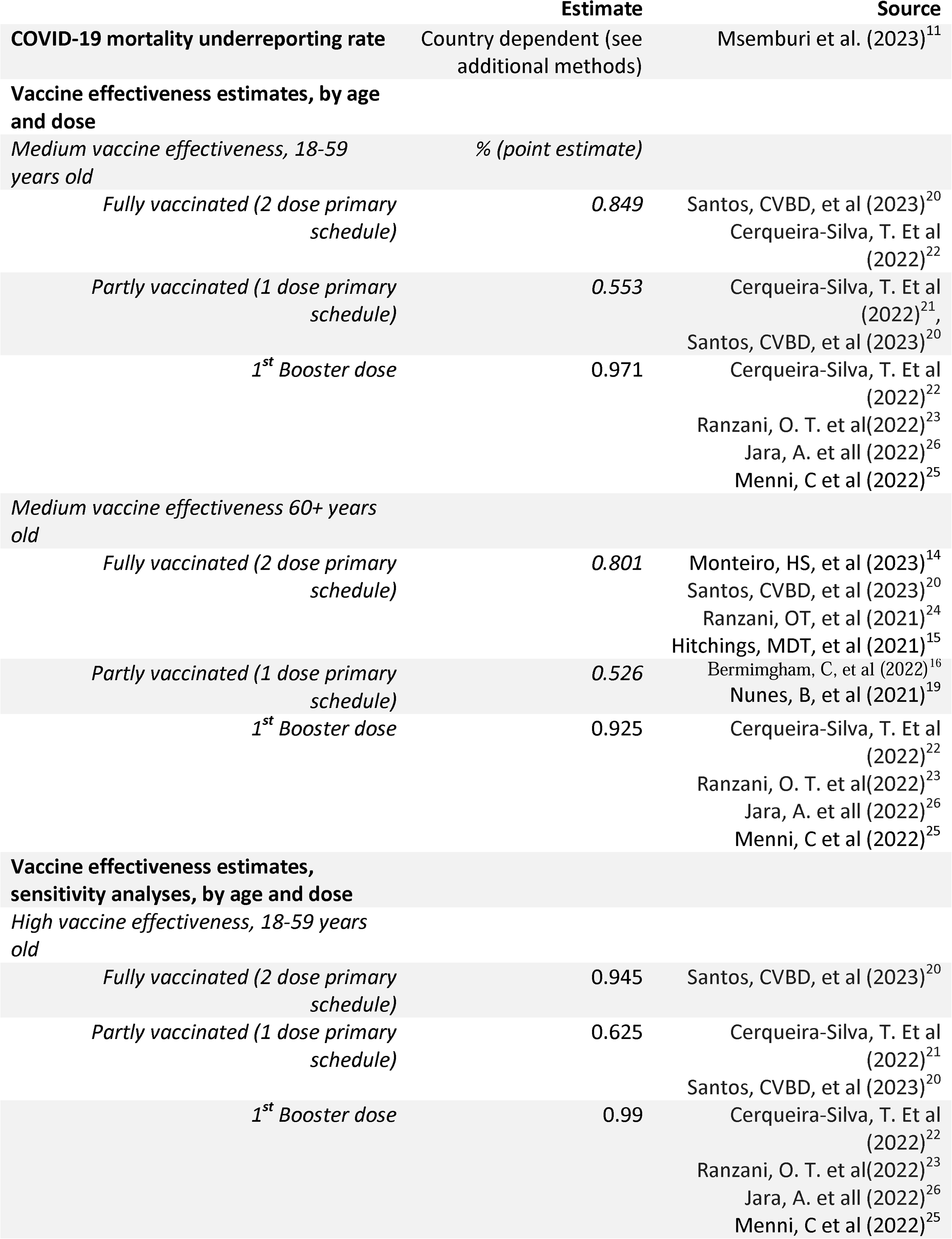
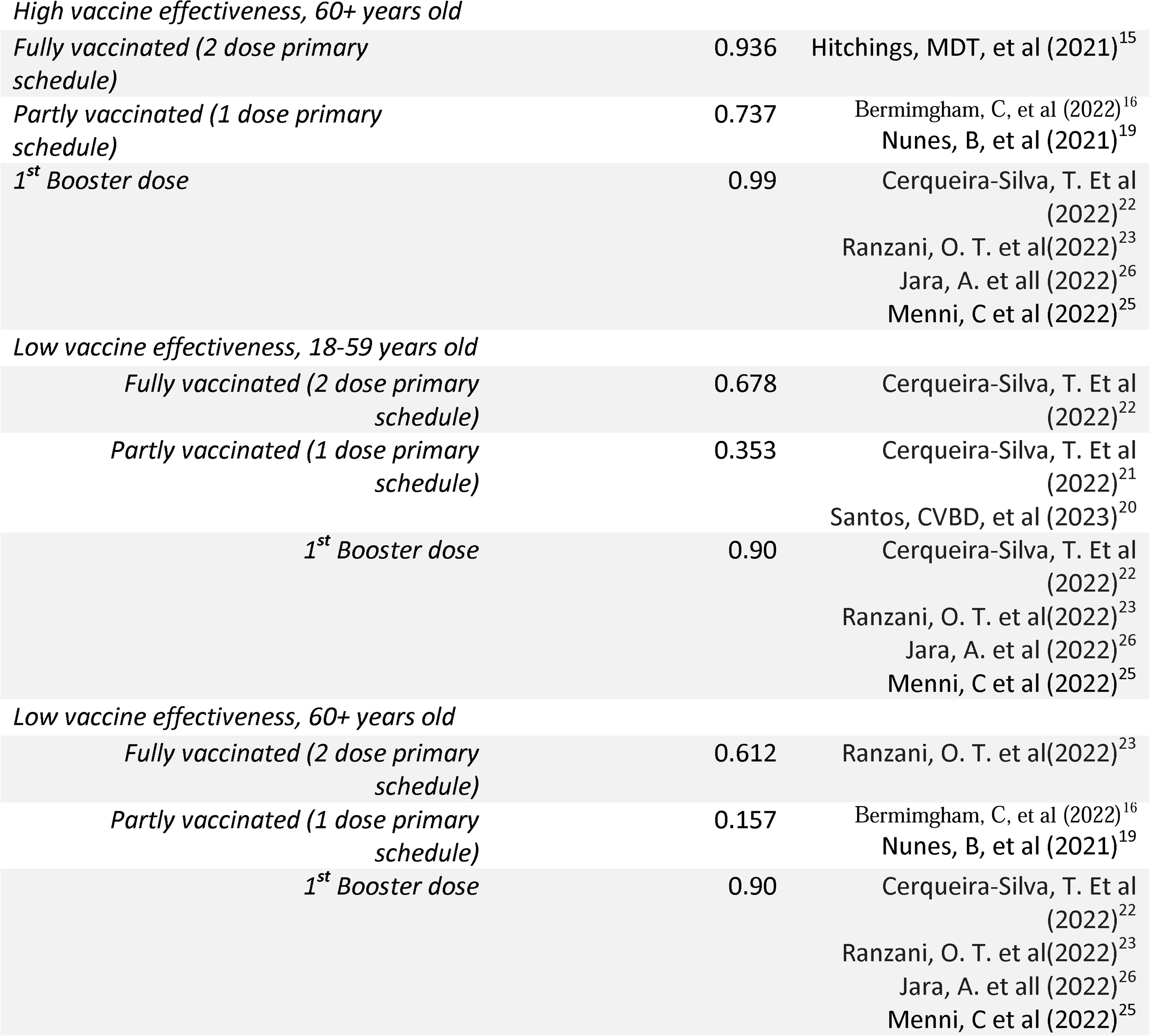
Parameters used as inputs to the model.

|  | Estimate | Source |
| --- | --- | --- |
| **COVID-19 mortality underreporting rate** | Country dependent (see additional methods) | Msemburi et al. (2023)<sup>11</sup> |
| **Vaccine effectiveness estimates, by age and dose** |  |  |
| *Medium vaccine effectiveness, 18-59 years old* | *% (point estimate)* |  |
| *Fully vaccinated (2 dose primary schedule)* | 0.849 | Santos, CVBD, et al (2023)<sup>20</sup><br>Cerqueira-Silva, T. Et al (2022)<sup>22</sup> |
| *Partly vaccinated (1 dose primary schedule)* | 0.553 | Cerqueira-Silva, T. Et al (2022)<sup>21</sup>,<br>Santos, CVBD, et al (2023)<sup>20</sup> |
| *1<sup>st</sup> Booster dose* | 0.971 | Cerqueira-Silva, T. Et al (2022)<sup>22</sup><br>Ranzani, O. T. et al(2022)<sup>23</sup><br>Jara, A. et all (2022)<sup>26</sup><br>Menni, C et al (2022)<sup>25</sup> |
| *Medium vaccine effectiveness 60+ years old* |  |  |
| *Fully vaccinated (2 dose primary schedule)* | 0.801 | Monteiro, HS, et al (2023)<sup>14</sup><br>Santos, CVBD, et al (2023)<sup>20</sup><br>Ranzani, OT, et al (2021)<sup>24</sup><br>Hitchings, MDT, et al (2021)<sup>15</sup> |
| *Partly vaccinated (1 dose primary schedule)* | 0.526 | Bermingham, C, et al (2022)<sup>16</sup><br>Nunes, B, et al (2021)<sup>19</sup> |
| *1<sup>st</sup> Booster dose* | 0.925 | Cerqueira-Silva, T. Et al (2022)<sup>22</sup><br>Ranzani, O. T. et al(2022)<sup>23</sup><br>Jara, A. et all (2022)<sup>26</sup><br>Menni, C et al (2022)<sup>25</sup> |
| **Vaccine effectiveness estimates, sensitivity analyses, by age and dose** |  |  |
| *High vaccine effectiveness, 18-59 years old* |  |  |
| *Fully vaccinated (2 dose primary schedule)* | 0.945 | Santos, CVBD, et al (2023)<sup>20</sup> |
| *Partly vaccinated (1 dose primary schedule)* | 0.625 | Cerqueira-Silva, T. Et al (2022)<sup>21</sup><br>Santos, CVBD, et al (2023)<sup>20</sup> |
| *1<sup>st</sup> Booster dose* | 0.99 | Cerqueira-Silva, T. Et al (2022)<sup>22</sup><br>Ranzani, O. T. et al(2022)<sup>23</sup><br>Jara, A. et all (2022)<sup>26</sup><br>Menni, C et al (2022)<sup>25</sup> |
| *High vaccine effectiveness, 60+ years old* |  |  |
| *Fully vaccinated (2 dose primary schedule)* | 0.936 | Hitchings, MDT, et al (2021)<sup>15</sup> |
| *Partly vaccinated (1 dose primary schedule)* | 0.737 | Bermingham, C, et al (2022)<sup>16</sup><br>Nunes, B, et al (2021)<sup>19</sup> |
| *1<sup>st</sup> Booster dose* | 0.99 | Cerqueira-Silva, T. Et al (2022)<sup>22</sup><br>Ranzani, O. T. et al(2022)<sup>23</sup><br>Jara, A. et all (2022)<sup>26</sup><br>Menni, C et al (2022)<sup>25</sup> |
| *Low vaccine effectiveness, 18-59 years old* |  |  |
| *Fully vaccinated (2 dose primary schedule)* | 0.678 | Cerqueira-Silva, T. Et al (2022)<sup>22</sup> |
| *Partly vaccinated (1 dose primary schedule)* | 0.353 | Cerqueira-Silva, T. Et al (2022)<sup>21</sup><br>Santos, CVBD, et al (2023)<sup>20</sup> |
| *1<sup>st</sup> Booster dose* | 0.90 | Cerqueira-Silva, T. Et al (2022)<sup>22</sup><br>Ranzani, O. T. et al(2022)<sup>23</sup><br>Jara, A. et al (2022)<sup>26</sup><br>Menni, C et al (2022)<sup>25</sup> |
| *Low vaccine effectiveness, 60+ years old* |  |  |
| *Fully vaccinated (2 dose primary schedule)* | 0.612 | Ranzani, O. T. et al(2022)<sup>23</sup> |
| *Partly vaccinated (1 dose primary schedule)* | 0.157 | Bermingham, C, et al (2022)<sup>16</sup><br>Nunes, B, et al (2021)<sup>19</sup> |
| *1<sup>st</sup> Booster dose* | 0.90 | Cerqueira-Silva, T. Et al (2022)<sup>22</sup><br>Ranzani, O. T. et al(2022)<sup>23</sup><br>Jara, A. et all (2022)<sup>26</sup><br>Menni, C et al (2022)<sup>25</sup> |

#### Population

Population estimates by age sub-group for each country in 2021 were obtained from the United Nations World Populations Prospects.^27^

### Model Structure and Equations

To calculate deaths averted via vaccination, we used the following equations, wherein ***D_i,m,V=v_*** is the number of deaths reported in month ***i***, for age group ***m***. The reported level of vaccination for age group ***m*** at month ***i*** is shown as ***v***. ***D_i,m,V=O_*** is the expected number of deaths that would have been observed for age group ***m*** at month ***i*** had there been no vaccination, and ***X*** is a multiplier for assumed underreporting of case counts.^18^ ***p_o,i,m_*** is the proportion of the population partly vaccinated (one dose), p_f,i,m_ is the proportion of the population fully vaccinated, and ***p_b,i,m_*** is the proportion of the population booster vaccinated for age group ***m*** at month ***i***. ***VE_o_***, is the vaccine effectiveness against death for partial vaccination, ***VE_f_*** is the vaccine effectiveness against death for full series vaccination, and ***VE_b_*** is the vaccine effectiveness against death for full series vaccination with booster. ***D_Averted_*** is the estimated number of deaths averted via vaccination. Equation 1 shows the calculation for expected deaths that would have been observed if there had been no vaccination in age group ***m*** in month ***i***.

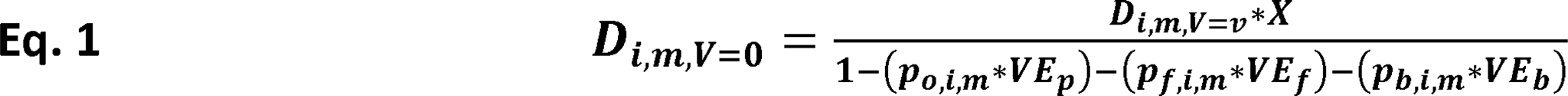

Equation 2 sums all months of observation for age group ***m***.

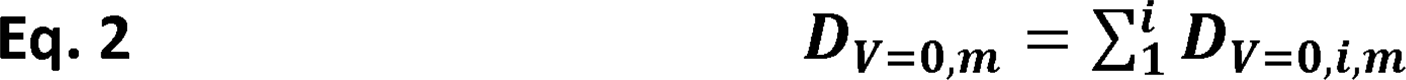

Equation 3 calculates estimated deaths averted via vaccination.

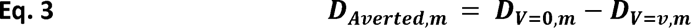

Equation 4 sums deaths averted across age groups for the final estimate of deaths averted.

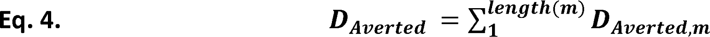

We calculated estimated deaths averted by country within the group of countries included in the regional analysis and used country-specific estimates to come to a final region-wide estimate.

### Sensitivity analysis

As there were multiple vaccine products used in this region over time, with varying estimates of vaccine effectiveness against COVID-19 due to different variants of concern (VOC), which also varies over time and by country, we considered a range of vaccine effectiveness estimates in the model, assigned to low, medium, and high range estimates. Also under sensitivity analysis, we modeled estimates with and without accounting for under-reporting of COVID-19 mortality.

### Ethical issues

This study used publicly available secondary data, considering a variety of open data sources and platforms which report COVID-19 related data reported and made available by the countries, PAHO, WHO and other international organizations. Only aggregated data were used, and mathematical models have been applied to estimate the averted death burden associated with COVID-19 vaccination in the region.

## Results

There were 1.05 million COVID-19 deaths reported in the 17 selected Latin America and Caribbean countries from the start of vaccination (ranging by country from December 2020 to March 2021) to May 2022 (Table 1). Accounting for underreporting, the analysis assumes that there were actually 1.49 million COVID-19 deaths in these countries during this period. (Table 3, Figure 2). Our model estimates that without vaccination and assuming medium vaccine effectiveness, there would have been 2.67 million deaths during this same time period. Therefore, an estimated 1.18 million deaths were averted by vaccination, with that estimate ranging between 610,000 deaths averted assuming low vaccine effectiveness and 2.62 million deaths averted assuming high vaccine effectiveness. Overall, our model estimates that around 273 (142-607) deaths were averted per 100,000 people in LAC.

**Figure 2.**
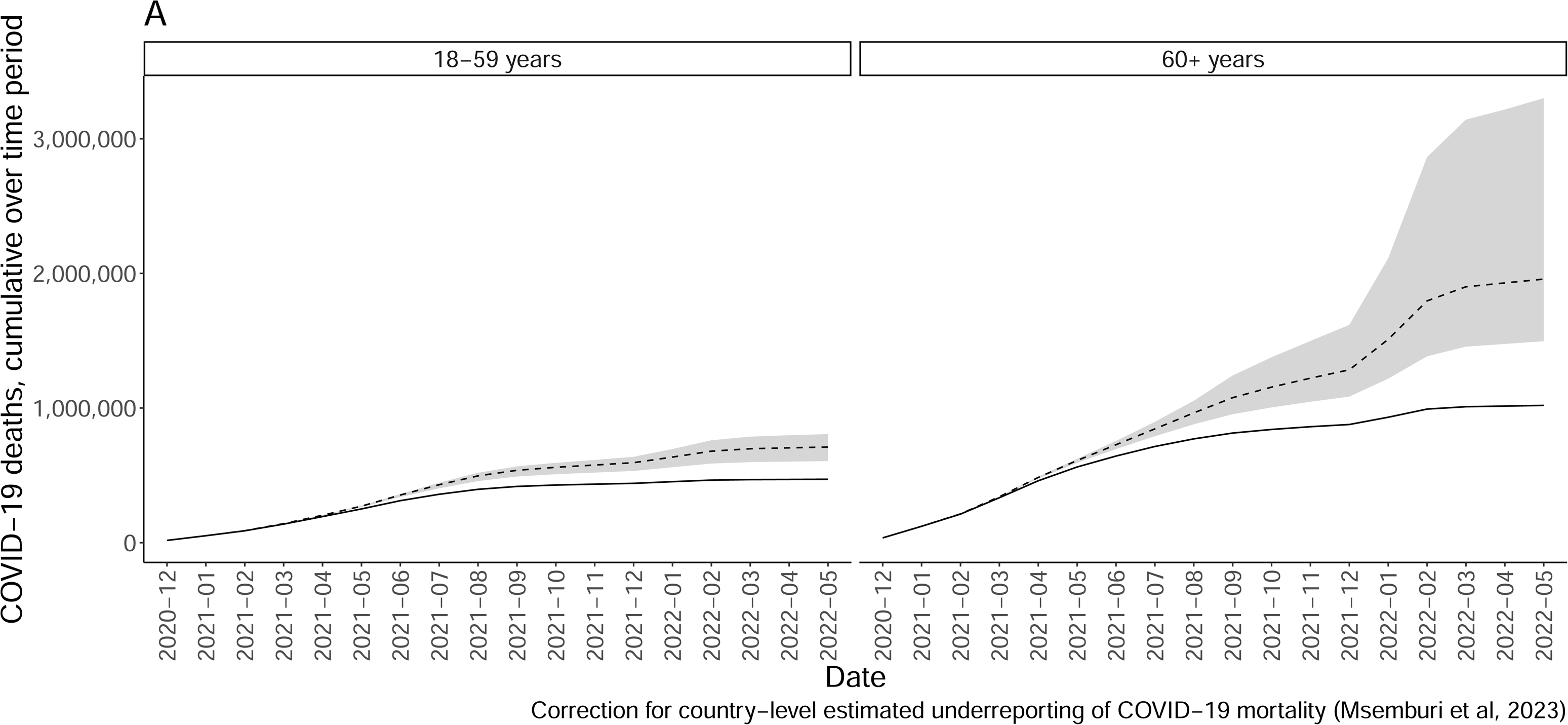
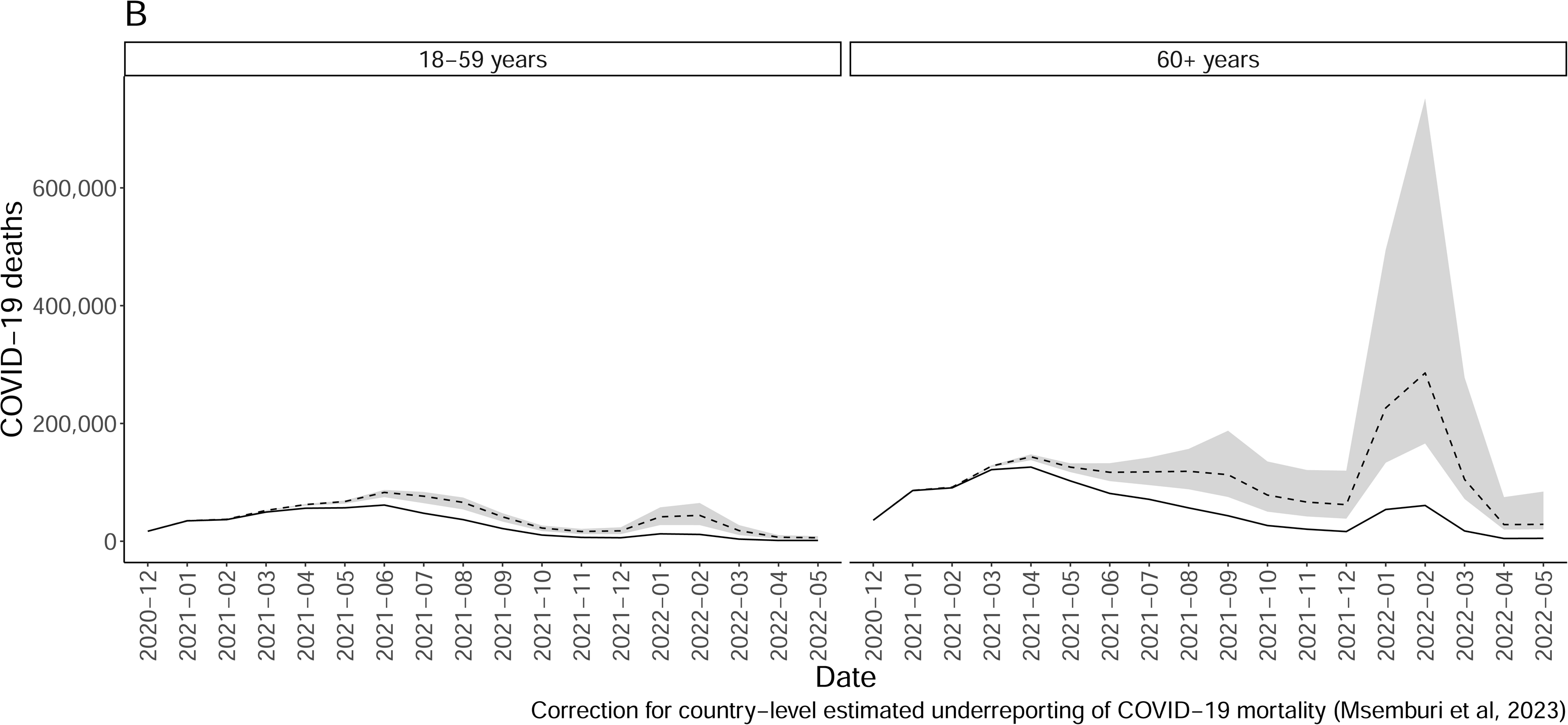
Observed deaths (solid line) and model estimates for deaths without vaccination (dashed line) by age-group (60+: blue, 18-59: red) over time. A. Cumulative deaths over time, B. Incident deaths over time. The shaded grey area represents the range of deaths averted given range of vaccine effectiveness estimates.

**Table 3.** Model estimates for deaths averted by COVID-19 vaccination, stratified by vaccine effectiveness ranges, with and without correction for under-reporting of deaths, during the study period. Selected countries in the Latin American and Caribbean Region.

|  |  | **Medium Vaccine Effectiveness** |  | **Low Vaccine Effectiveness** |  | **High Vaccine Effectiveness** |  |
| --- | --- | --- | --- | --- | --- | --- | --- |
|  | *Observed deaths* | *Estimated number of deaths, no vaccination* | *Estimated deaths averted* | *Estimated number of deaths, no vaccination* | *Estimated deaths averted* | *Estimated number of deaths, no vaccination* | *Estimated deaths averted* |
| **Region estimates** |  |  |  |  |  |  |  |
| *Correction for underreporting* | 1,490,000 | 2,670,000 | 1,180,000 | 2,100,000 | 610,000 | 4,110,000 | 2,620,000 |
| *No correction for underreporting* | 1,050,000 | 1,920,000 | 870,000 | 1,500,000 | 450,000 | 3,010,000 | 1,960,000 |

In a sensitivity analysis, if we instead considered COVID-19 deaths as reported with no assumed underreporting, our model estimated that vaccination averted 870,000 deaths (range: 450,000, 1.96 million) (Table 3, Supplementary Figure 2)

Country-specific data closely align with region-wide results (Supplementary Table 1, Supplementary Figures 2,3). Rates of deaths averted by country varied considerably by country over time, especially in the over 60 age group (Supplemental Figure 4).

## Discussion

The COVID-19 pandemic has caused unprecedented loss of life worldwide, and vaccination has emerged as a critical tool to mitigate its impact. To our knowledge, this is the first multi-country study evaluating the impact of vaccination on COVID-19 deaths in the Latin American and Caribbean Region. Our model estimates that between 700,000 and 3.1 million deaths were averted by vaccination, underscoring the importance that vaccination had in mitigating the impact of the COVID-19 pandemic.

Incidence of deaths averted by country varied based on vaccination coverage over time (especially in the over 60 age group) (Supplementary Figure 4) as well as whether COVID-19 outbreaks were seen largely prior to or after the initiation of vaccination. For instance, the model estimates that proportionally more deaths were averted in Chile and Uruguay as there were spikes in deaths at a time when a lot of the population (especially those over 60) was vaccinated (Supplementary Figures 2,3).

Our results are in line with available model projections of COVID-19 deaths averted by vaccination in other Regions and in selected countries. The projected impact of COVID-19 vaccination in deaths during the first year of COVID-19 vaccination rollout globally during the first year of vaccine availability from Dec 8 2020 through Dec 8, 2021 was reported as 14·4 million (95% credible interval [Crl] 13·7-15·9) in 185 countries.^28^

Savinkina et al^8^ also modeled COVID-19 deaths-averted by vaccinating low-income and lower-middle-income countries in late 2021, when the Omicron VOC was already predominant globally. The study estimated that scaling up global vaccination in 2022 with complete primary series plus a booster dose could result in an additional 1.5 million COVID-19 deaths averted in low and lower-middle-income countries.

Country and region-specific studies have also been conducted. In the United States, Steele et al.^7^ modeled the projected number of deaths averted by COVID-19 vaccination, and concluded that the US vaccination campaign averted 58% of deaths that might have otherwise occurred in the period of Dec 1^st^ 2020 through September 30^th^, 2021. Over the same time period, our analysis estimates that approximately 25% (range based on vaccine effectiveness estimates: 15%-34%) of deaths had been averted via vaccination in LAC. When expanding the time frame through May 2022, our analysis estimates that an approximate 45% (range based on vaccine effectiveness estimates: 30%-65%) of deaths had been averted via vaccination. This may be due to vaccination starting earlier and being more readily and equitably accessible to the population in the US than in much of LAC.

Ferreira et al^29^ estimated averted COVID-19 deaths by vaccination in Brazil among adults over 60 years of age considering a time period from January to August 2021, reporting 58,000 averted deaths. The authors reinforce that this represents an underestimate of the impact of vaccination in Brazilian elderly. In this time period, our model estimates approximately 70,000 deaths averted in those over 60 years of age (range: 47,000-90,000).

A similar assessment for 33 countries of the European Region from December 2020, when vaccination was first introduced, through November 2021, resulted in an estimated 469,186 deaths averted (95% credible interval [Crl] 129,851–733,744; 23–62%).^30^

Despite varying geographical locations and scope, and different study periods, all these studies reinforce the significant impact of averted COVID-19 mortality due to vaccination, with varying rates by country, period, and vaccine coverage.

Our study has several limitations. A number of vaccine products were used in the countries in the analysis over the time-period of interest, but we did not have adequate data on vaccine coverage by each vaccine product.

Therefore, we averaged vaccine effectiveness across products and present three estimates, ranging from low vaccine effectiveness equal to the lowest effectiveness vaccine used in the region to high vaccine effectiveness, equal to the effectiveness of mRNA vaccines. Due to uncertainty in vaccine effectiveness, we did not consider vaccine waning in the analysis as we provide a wide range of vaccine effectiveness estimates that would capture vaccine waning. Our analysis relies on publicly available aggregated data as reported by countries, and some of this reporting may be incomplete due to the nature of data reporting during a pandemic. Some of this limitation, particularly related to under-reporting of COVID-19 mortality, was addressed by considering an adjustment factor for under-reporting. In addition, our analysis does not account for protection derived from past SARS-CoV2 natural infection against reinfection and severe disease.^31^ This could result in a potential over-estimation of vaccine impact on deaths in our case-base scenario. This is somewhat accounted for in sensitivity analysis considering a range of estimates for potential vaccine effect. Lastly, we did not use a dynamic transmission model, and therefore cannot model disease dynamics in the population over time nor the indirect protection resulting from vaccination. Considering the reported indirect effects of the vaccine in decreasing the probability of secondary transmission from an infected vaccine recipient to^32^ other individual, potentially resulting in an even more significant impact of the vaccine in terms of reducing deaths, our estimates are conservative and would be underestimating vaccine impact.

This analysis captured data from 17 countries, for which data were available. Though there are more countries in Latin America and the Caribbean, the countries excluded from the analysis were mainly small and would not greatly change the magnitude of the overall estimated results. However, these countries were largely low income with high burden of COVID-19 disease; it is important to continue working to improve data collection and surveillance in this region in order to make analyses possible in the future. Also, our model does not take into account other public health measures implemented by the countries during the pandemic, and adherence to them by the population, including mask use, lockdowns, quality and availability of healthcare services, among others. Though these factors may impact COVID-19 mortality, but it was not possible to ascertain their individual impact in this modelling study.

Finally, it is worthwhile mentioning that such analysis is only made possible due to the availability of administrative data collected and shared by countries. During a pandemic and in public health emergency situations, data collection, cleaning, sharing and analysis may be even more challenging and pose an additional burden to already constrained public health healthcare personnel in the local levels. However, analyses such as this one underscore the importance of data collection and analysis to support decision making, public health policy implementation, and assessment of interventions.

In conclusion, our study provides evidence of the significant impact of vaccination in reducing COVID-19 deaths in the Latin American and Caribbean region, one of the regions most strongly impacted by the pandemic. Despite the many challenges to COVID-19 vaccination in LAC- including timely access to vaccines, varying vaccine products and schedules, evolving circulating variants, and shifting vaccination strategies and target groups-these findings underscore the underscore the substantial impact of timely and widespread vaccination in averting COVID-19 deaths. Further studies evaluating the impact of vaccination in other selected health outcomes including hospitalization, healthcare service utilization, costs, and long-COVID, in addition to other non-health outcomes including educational, social and economic indicators, may provide additional evidence relevant to policy makers and society as a whole. All these constitute complementary pieces of evidence which are valuable support for vaccination programs aimed at combating epidemic infectious diseases in the region and future pandemics.

## Data Sharing Statement

All data used in this analysis are publicly available. All analysis code is publicly available and can be accessed at: https://github.com/ASavinkina/COVID_Vx_Impact.

## Presentation

This work was previously presented at Epidemics, in November 2023.

## Data Availability

All data produced are available online at

https://github.com/ASavinkina/COVID_Vx_Impact

## Acknowledgement

We are thankful to the PAHO team who shared immunization coverage data, and teams responsible for data collection, cleaning and sharing in regional and global data repositories.

---

**Supplementary Figure 1.** Observed deaths (solid line) and model estimates for deaths without vaccination (dashed line) by age-group (18-59: left, 60+: right panel) over time. A. Cumulative deaths over time, B. Incident deaths over time. No correction for underreporting of COVID-19 deaths.

**Supplementary Figure 2.** Incident observed deaths, per 100,000 population, (solid line) and model estimates for deaths without vaccination, per 100,000, (dashed line) by age-group (18-59: left, 60+: right panel) over time, by country.

**Supplementary Figure 3.** Incident observed deaths, per 100,000 population, (solid line) and model estimates for deaths without vaccination, per 100,000, (dashed line) by age-group (18-59: left, 60+: right panel) over time, by country. No correction for underreporting of COVID-19 deaths.

**Supplementary Figure 4.** Incident deaths averted per 100,000 population (vertical axis) plotted against vaccination coverage (two dose vaccination) (horizontal axis). A. Under 60 years of age, B. Over 60 years of age.

