## Additional Methods for "Deaths averted by COVID-19 vaccination in select Latin American and Caribbean Countries: a modelling study"

***Deaths by age imputation***

COVID-19 death data by age (18-59, 60+) from the COVerAGE-DB database was unavailable for the following countries: Belize, Bolivia, Costa Rica, Ecuador, El Salvador, Guatemala, Honduras, and Venezuela. Deaths data for the time period in question was available from the WHO, however this data was not age-stratified. We therefore imputed proportion over deaths by age group for these countries using a linear regression, building the prediction model using data from countries with complete age-startified COVID-19 deaths data from COVerAGE-DB.

Predictors used in the model were month and year, proportion of population under 60 years of age, and the proportion of the population over 60 who are vaccinated (partly and fully). Outcome predicted was proportion of deaths in those over 60. The prediction model was validated by individually being run on each country with full data, and then comparing observed proportions of death in those over 60 to predicted proportions of death in those over 60. See Table below for observed and predicted values for each country.

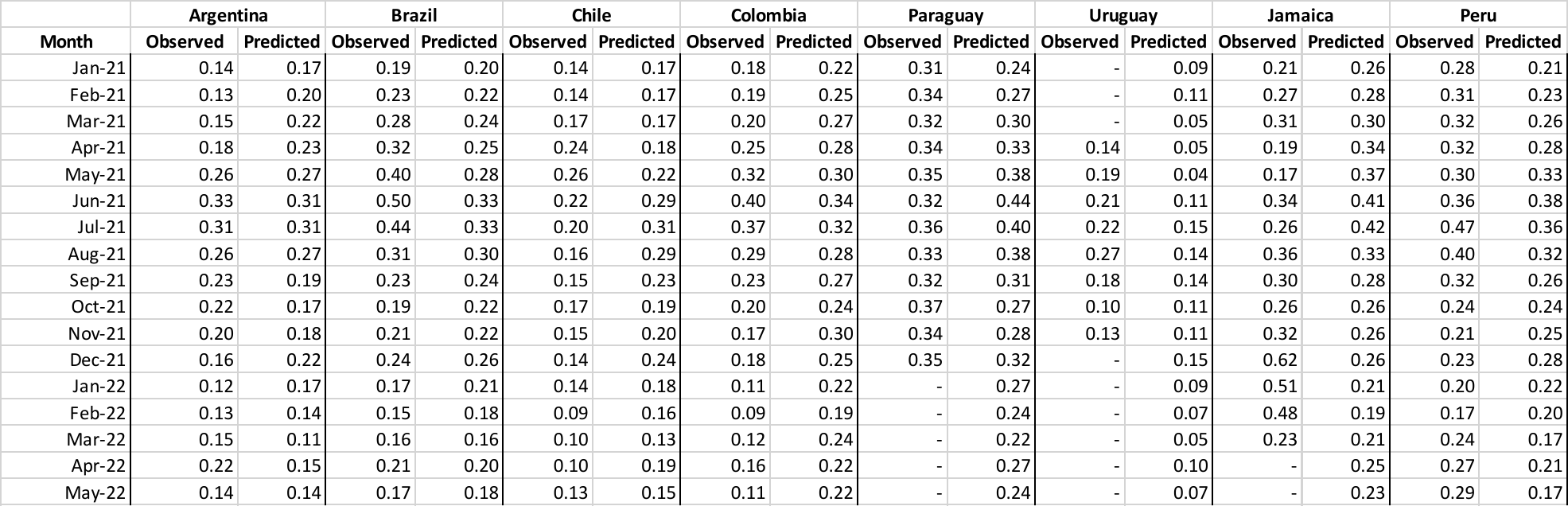

Predicted values for proportion of deaths in those over 60 for countries with no data, as well as for countries with missing data (Paraguay, Uruguay, Jamaica) were then applied to the number of deaths observed in that month in order to divide deaths by age group.

***National Underreporting Estimates***

National underreporting estimates are taken from Msemburi et. al 2023.^1^ To get an underreporting estimate, we compared reported number of COVID-19 deaths in 2021 to the number of excess deaths in the country during the same time frame. If the number of COVID-19 deaths exceeded excesss deaths, we assumed there was no underreporting.

| Country | ISO3 | Year | Reported COVID Deaths | Excess Deaths | Underreporting |
| --- | --- | --- | --- | --- | --- |
| Antigua and Barbuda | ATG | 2021 | 113 | 27.07 | -3.1743628 |
| Argentina | ARG | 2021 | 74093 | 58190.5 | -0.2732834 |
| Bahamas | BHS | 2021 | 546 | 667.44 | 0.18194894 |
| Barbados | BRB | 2021 | 253 | -95.95 | 3.63679 |
| Belize | BLZ | 2021 | 356 | 574.98 | 0.38084803 |
| Bolivia (Plurinational State of) | BOL | 2021 | 10515 | 51731.99 | 0.79674086 |
| Brazil | BRA | 2021 | 426136 | 470456.44 | 0.09420732 |
| Chile | CHL | 2021 | 22597 | 24119.93 | 0.0631399 |
| Colombia | COL | 2021 | 87246 | 110683.86 | 0.21175499 |
| Costa Rica | CRI | 2021 | 5198 | 8290.03 | 0.37298176 |
| Cuba | CUB | 2021 | 8177 | 18393.56 | 0.55544223 |
| Dominica | DMA | 2021 | 45 | 69.64 | 0.35381964 |
| Dominican Republic | DOM | 2021 | 1837 | 10797.87 | 0.82987386 |
| Ecuador | ECU | 2021 | 19646 | 34465.67 | 0.42998352 |
| El Salvador | SLV | 2021 | 2496 | 9529.02 | 0.73806331 |
| Grenada | GRD | 2021 | 199 | -13.68 | 15.5467836 |
| Guatemala | GTM | 2021 | 11299 | 40647.14 | 0.72202226 |
| Guyana | GUY | 2021 | 887 | 2415.5 | 0.63278824 |
| Haiti | HTI | 2021 | 530 | 6743.71 | 0.92140825 |
| Honduras | HND | 2021 | 7323 | 15377.4 | 0.52378165 |
| Jamaica | JAM | 2021 | 2168 | 4494.59 | 0.51764232 |
| Mexico | MEX | 2021 | 155119 | 311676.27 | 0.50230731 |
| Nicaragua | NIC | 2021 | 52 | 2928.99 | 0.98224644 |
| Panama | PAN | 2021 | 3492 | 4645.13 | 0.24824494 |
| Paraguay | PRY | 2021 | 14404 | 17406.87 | 0.17251062 |
| Peru | PER | 2021 | 109518 | 155851.18 | 0.29729117 |
| Saint Kitts and Nevis | KNA | 2021 | 28 | -76.35 | 1.36673216 |
| Saint Lucia | LCA | 2021 | 290 | 479.91 | 0.39572003 |
| Saint Vincent and the Grenadines | VCT | 2021 | 83 | 386.55 | 0.78528004 |
| Suriname | SUR | 2021 | 1069 | 1013.26 | -0.0550106 |
| Trinidad and Tobago | TTO | 2021 | 2699 | 1938.91 | -0.3920192 |
| Uruguay | URY | 2021 | 6000 | 5351.6 | -0.12116 |
| Venezuela (Bolivarian Republic of) | VEN | 2021 | 4300 | 18683.91 | 0.76985545 |

1.	Msemburi W, Karlinsky A, Knutson V, Aleshin-Guendel S, Chatterji S, Wakefield J. The WHO estimates of excess mortality associated with the COVID-19 pandemic. *Nature* 2023; **613**(7942): 130-7.
