## Supplementary figures and images for "Deaths averted by COVID-19 vaccination in select Latin American and Caribbean Countries: a modelling study"

### Supplemental Figure 1

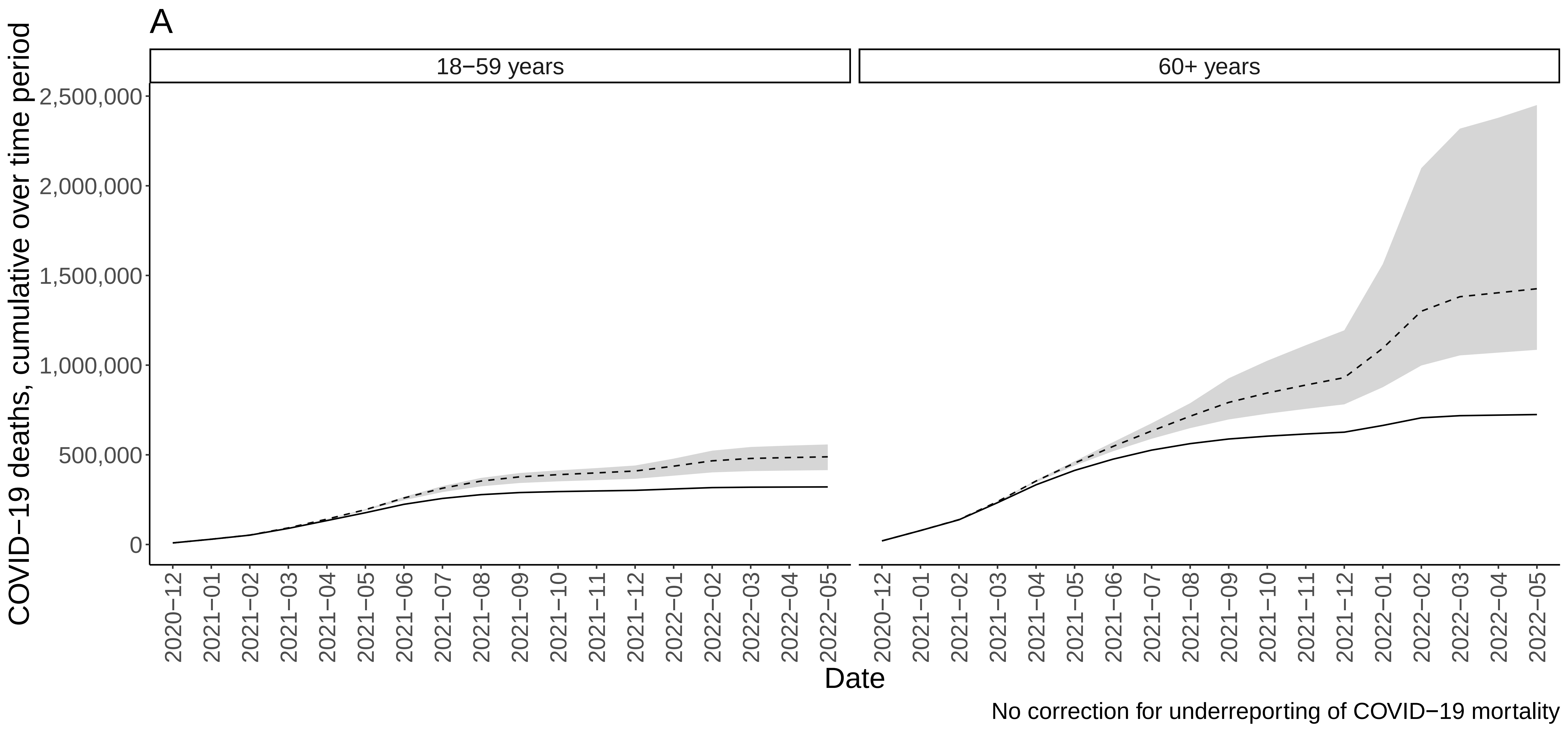

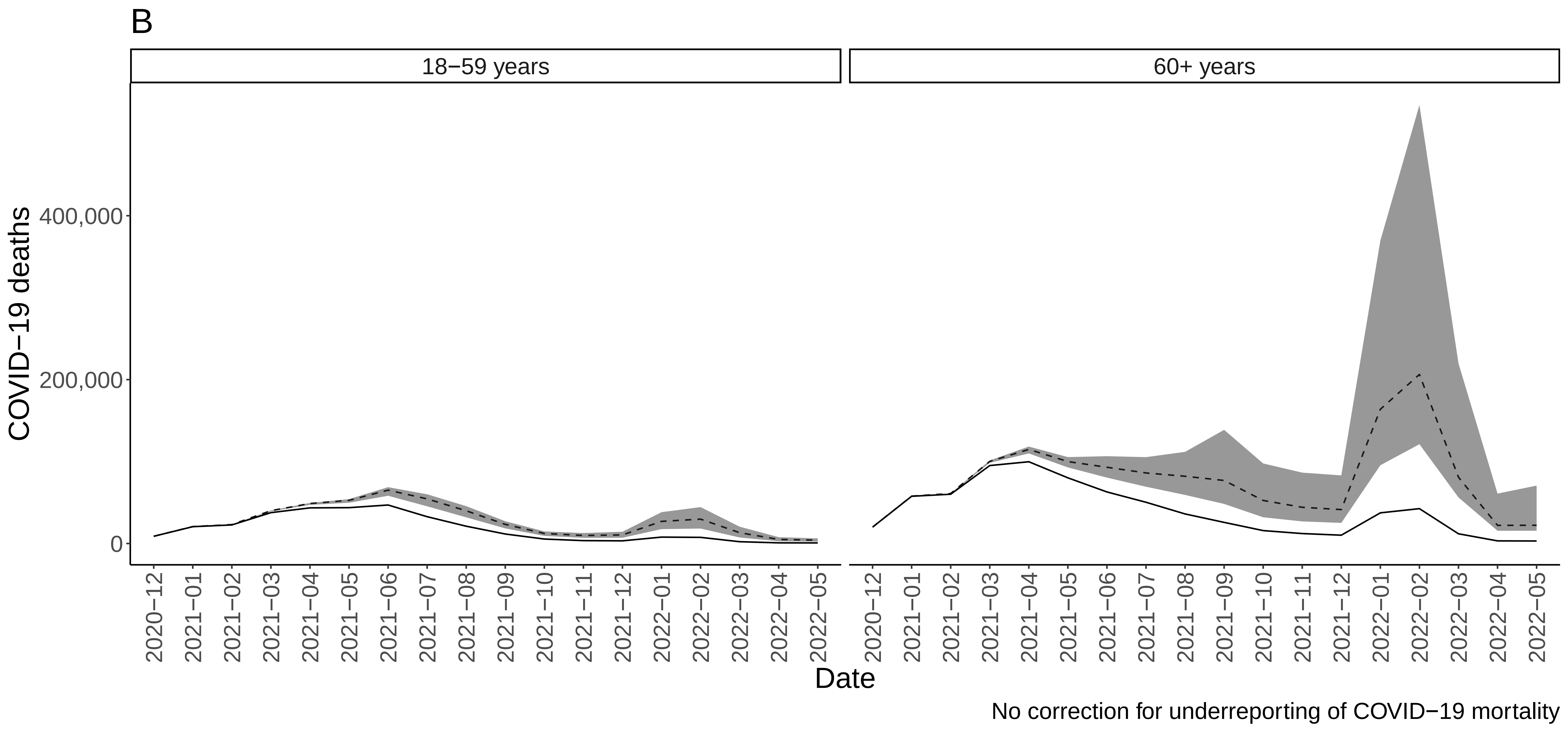

### Supplemental Figure 4

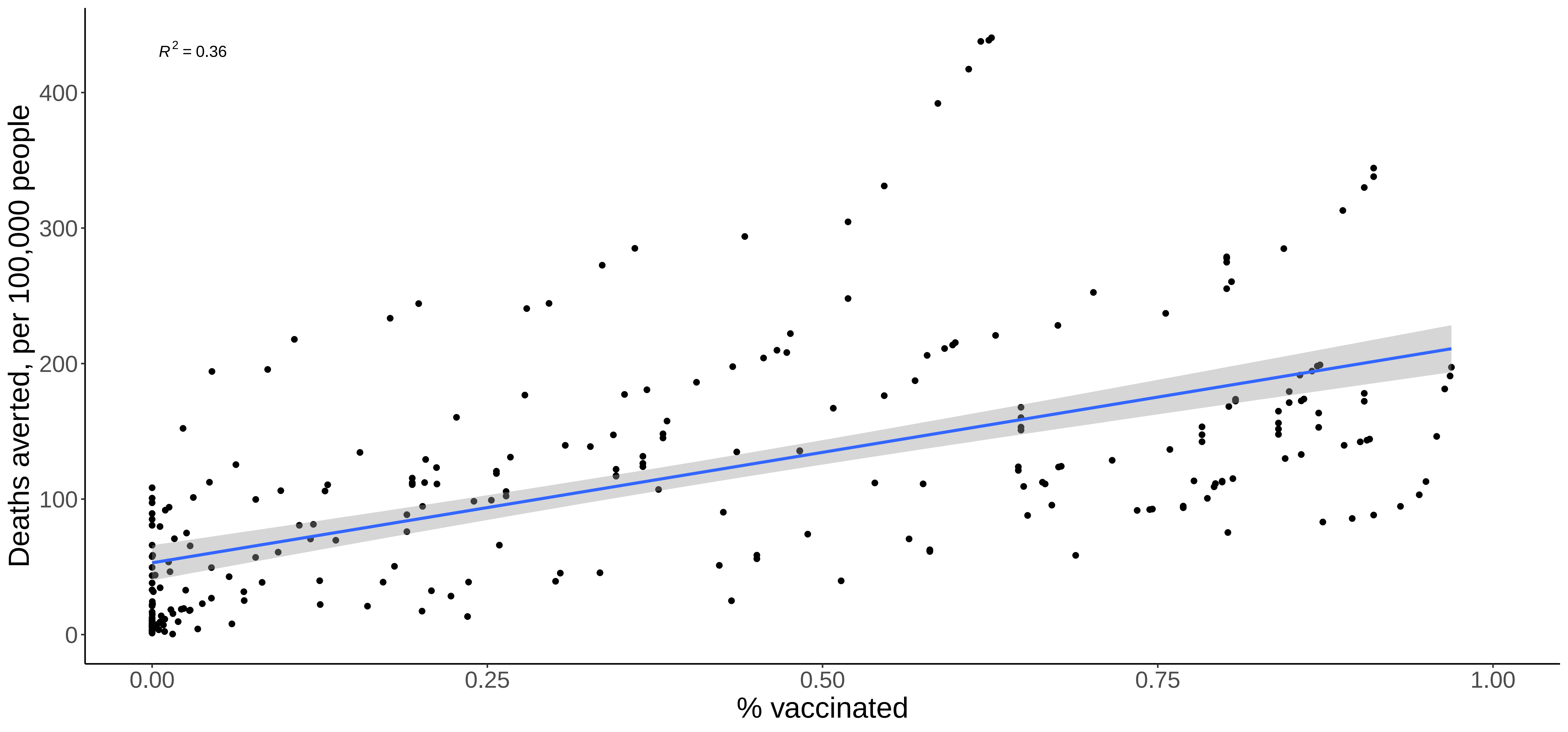

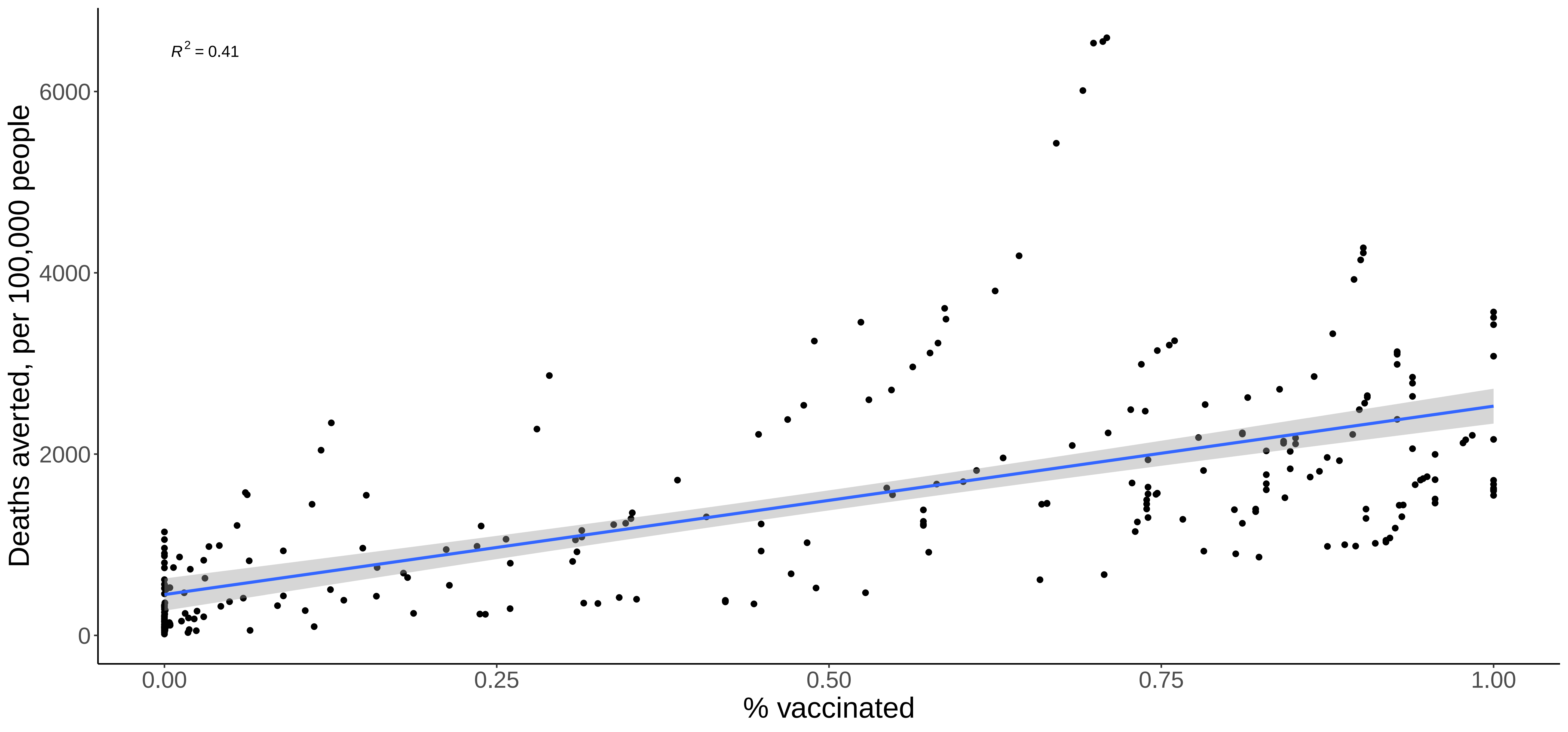
