## Supplemental Figure 2 for "Deaths averted by COVID-19 vaccination in select Latin American and Caribbean Countries: a modelling study"

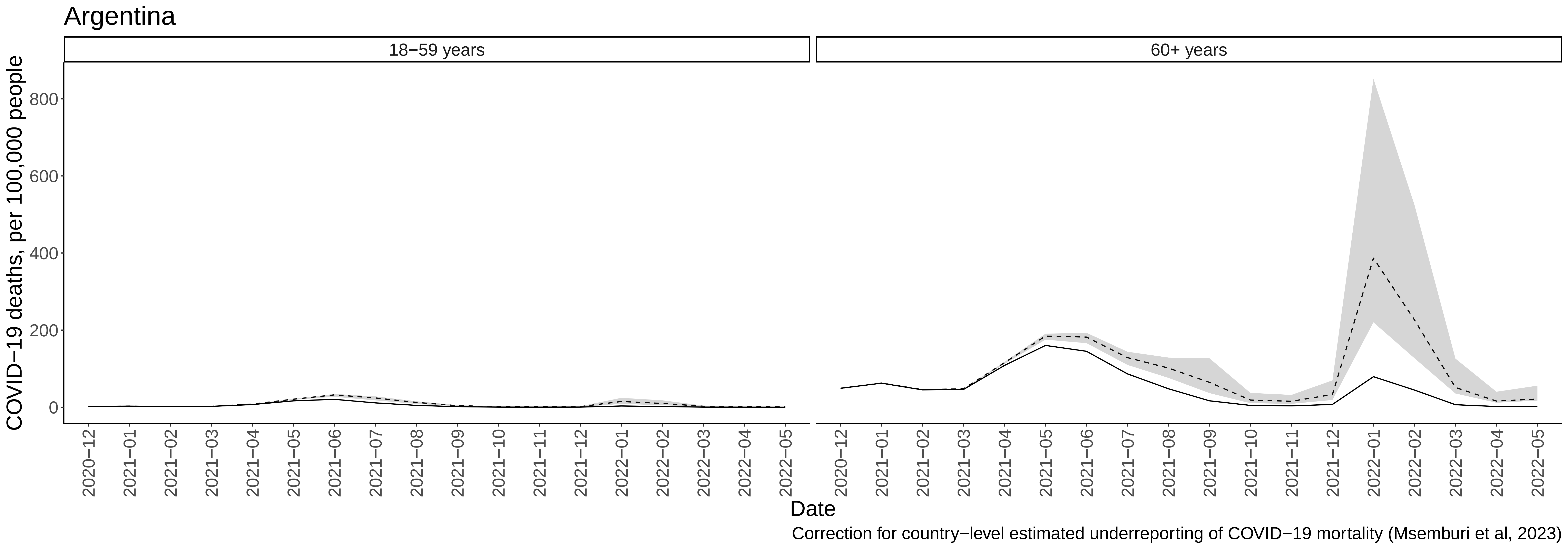
Argentina
18−59 years
60+ years
COVID−19 deaths, per 100,000 people
800
600
400
200
0
2020−12
2021−01
2021−02
2021−03
2021−04
2021−05
2021−06
2021−07
2021−08
2021−09
2021−10
2021−11
2021−12
2022−01
2022−02
2022−03
2022−04
2022−05
Date
Correction for country−level estimated underreporting of COVID−19 mortality (Msemburi et al, 2023)

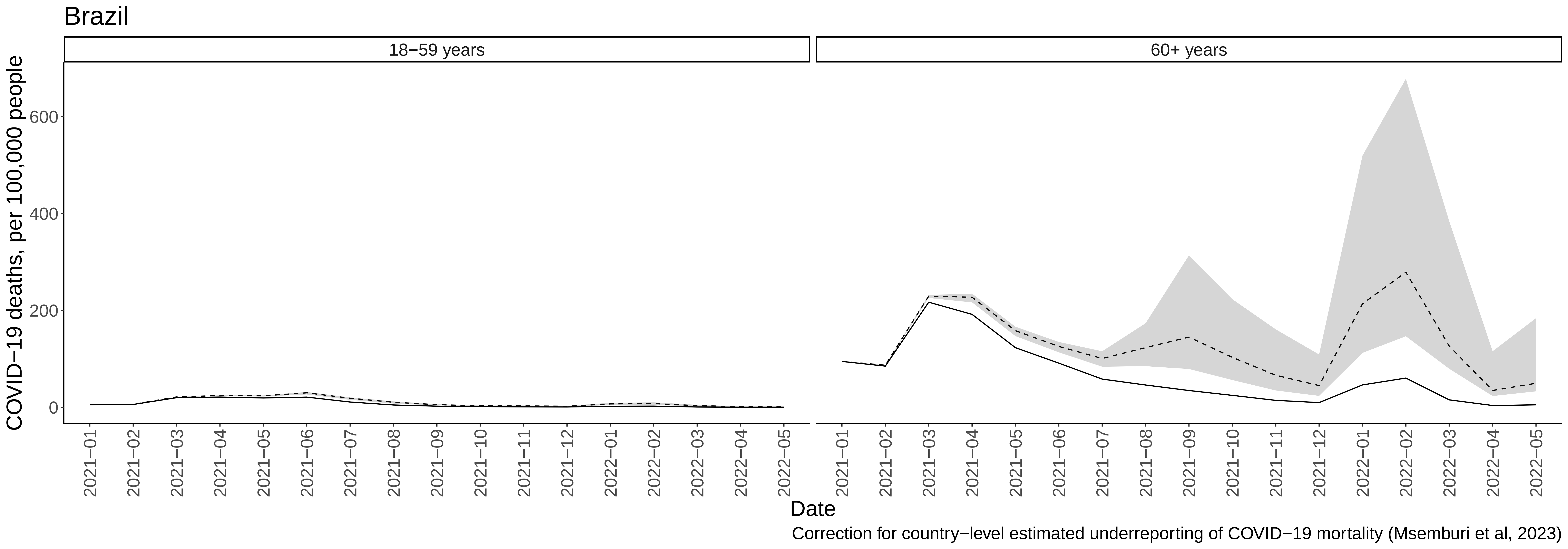

Brazil
18−59 years
60+ years
COVID−19 deaths, per 100,000 people
0
200
400
600
2021−01
2021−02
2021−03
2021−04
2021−05
2021−06
2021−07
2021−08
2021−09
2021−10
2021−11
2021−12
2022−01
2022−02
2022−03
2022−04
2022−05
Date
Correction for country−level estimated underreporting of COVID−19 mortality (Msemburi et al, 2023)

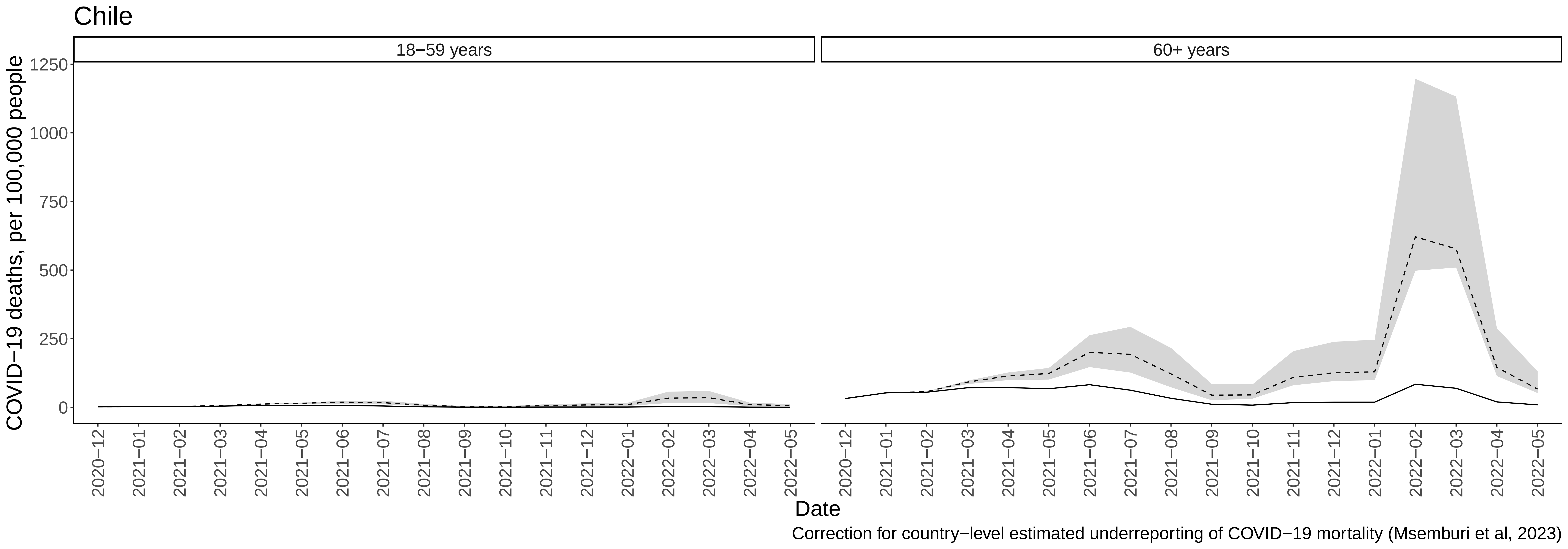
Chile
18−59 years
60+ years
COVID−19 deaths, per 100,000 people
1250
1000
750
500
250
0
2020−12
2021−01
2021−02
2021−03
2021−04
2021−05
2021−06
2021−07
2021−08
2021−09
2021−10
2021−11
2021−12
2022−01
2022−02
2022−03
2022−04
2022−05
Date
Correction for country−level estimated underreporting of COVID−19 mortality (Msemburi et al, 2023)

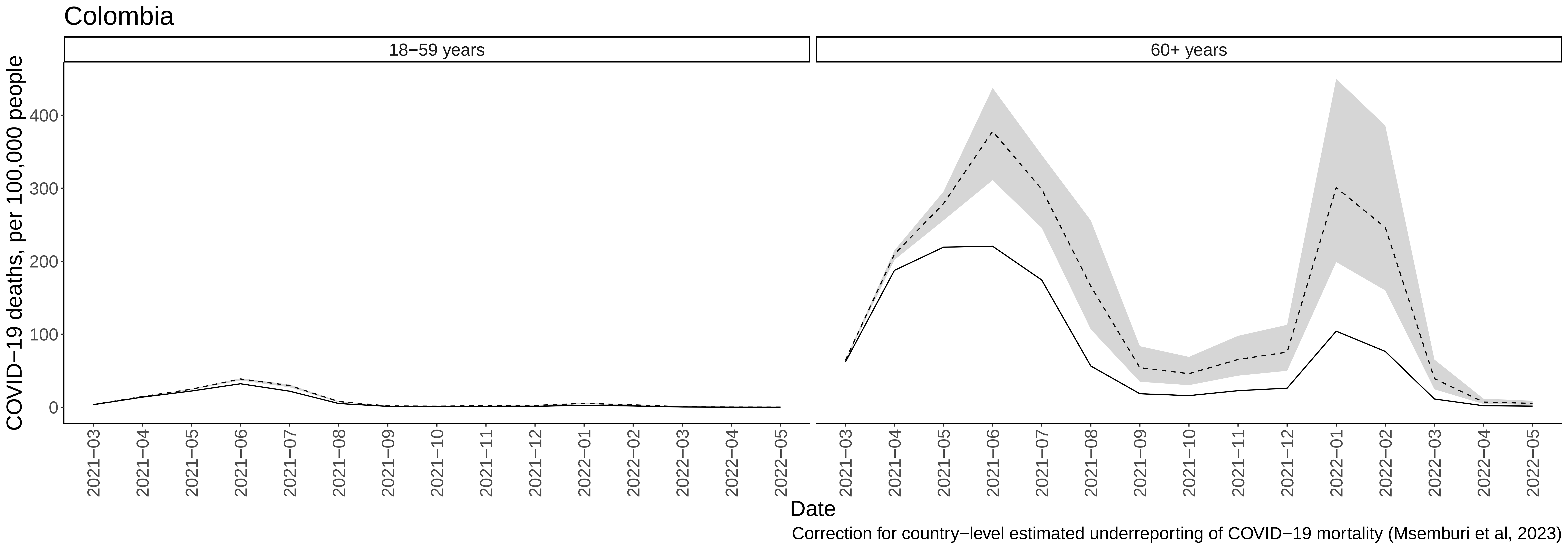

Colombia
18−59 years
60+ years
COVID−19 deaths, per 100,000 people
0
100
200
300
400
2021−03
2021−04
2021−05
2021−06
2021−07
2021−08
2021−09
2021−10
2021−11
2021−12
2022−01
2022−02
2022−03
2022−04
2022−05
Date
Correction for country−level estimated underreporting of COVID−19 mortality (Msemburi et al, 2023)

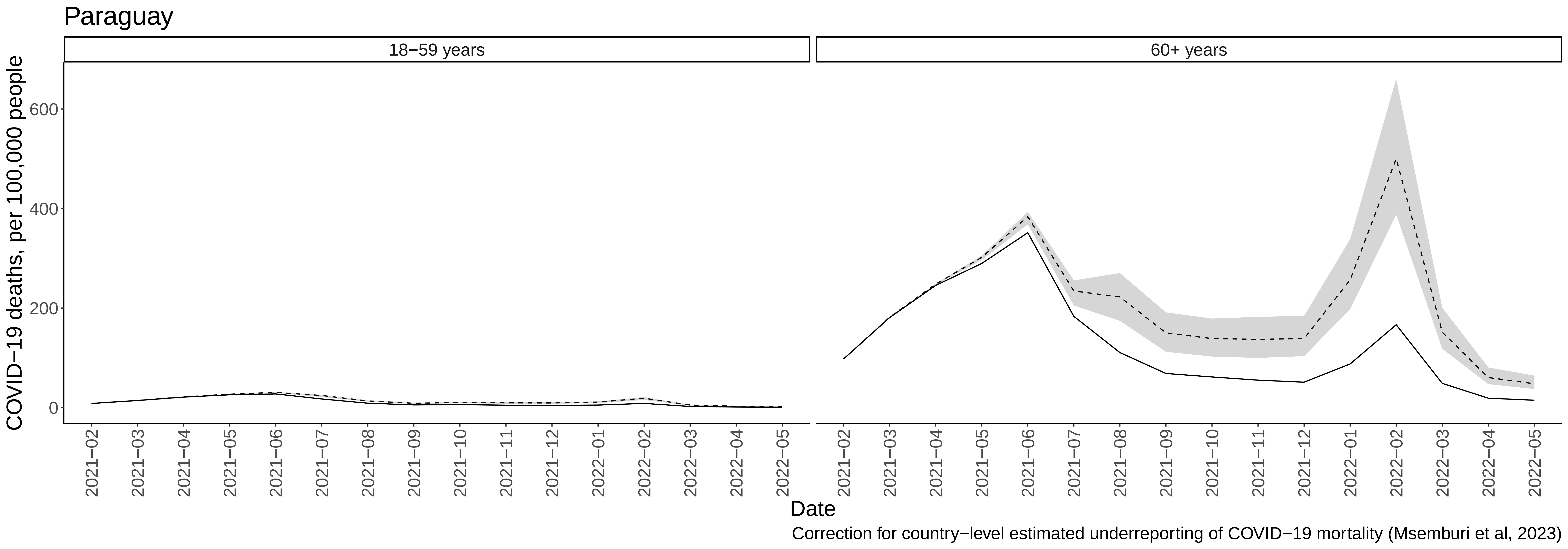

Paraguay
18−59 years
60+ years
COVID−19 deaths, per 100,000 people
0
200
400
600
2021−02
2021−03
2021−04
2021−05
2021−06
2021−07
2021−08
2021−09
2021−10
2021−11
2021−12
2022−01
2022−02
2022−03
2022−04
2022−05
Date
Correction for country−level estimated underreporting of COVID−19 mortality (Msemburi et al, 2023)

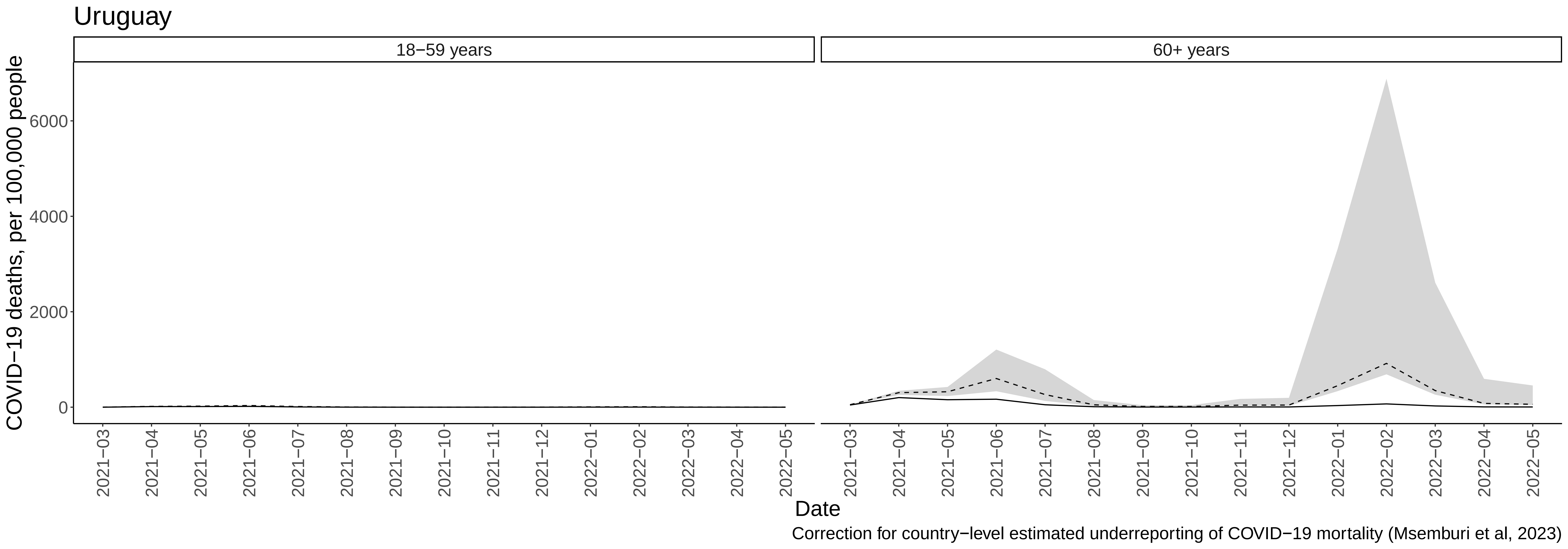

Uruguay
18−59 years
60+ years
COVID−19 deaths, per 100,000 people
0
2000
4000
6000
2021−03
2021−04
2021−05
2021−06
2021−07
2021−08
2021−09
2021−10
2021−11
2021−12
2022−01
2022−02
2022−03
2022−04
2022−05
Date
Correction for country−level estimated underreporting of COVID−19 mortality (Msemburi et al, 2023)

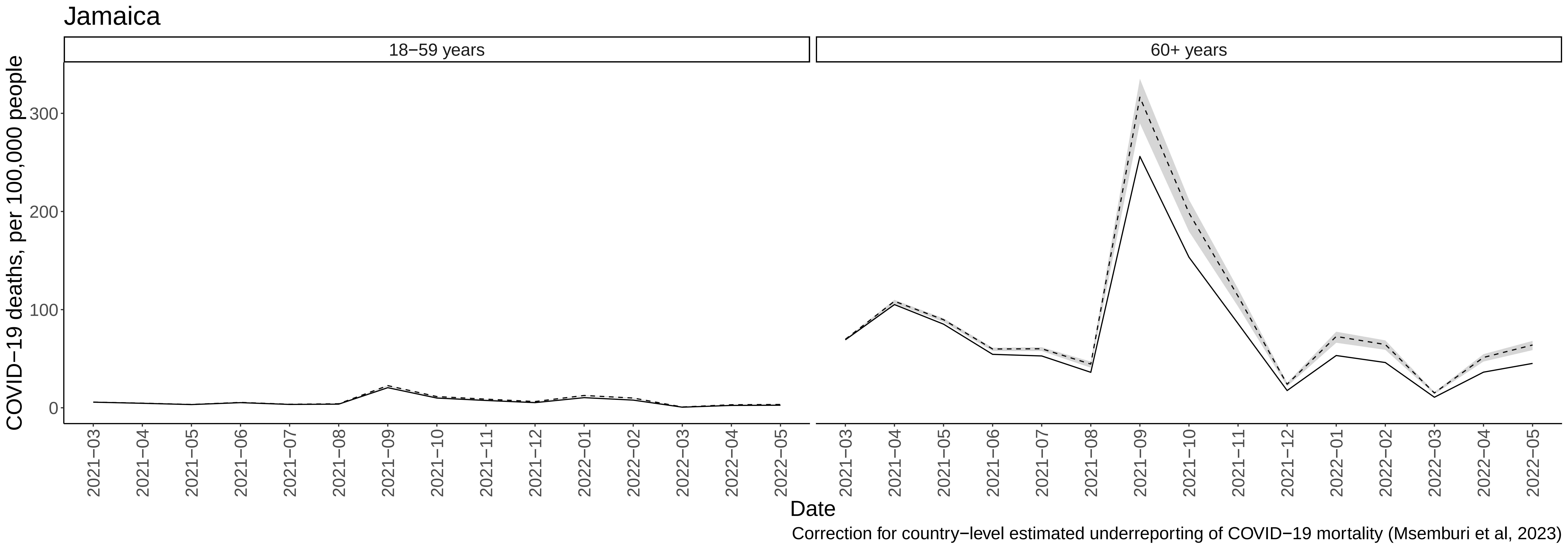
Jamaica
18−59 years
60+ years
COVID−19 deaths, per 100,000 people
0
100
200
300
2021−03
2021−04
2021−05
2021−06
2021−07
2021−08
2021−09
2021−10
2021−11
2021−12
2022−01
2022−02
2022−03
2022−04
2022−05
Date
Correction for country−level estimated underreporting of COVID−19 mortality (Msemburi et al, 2023)

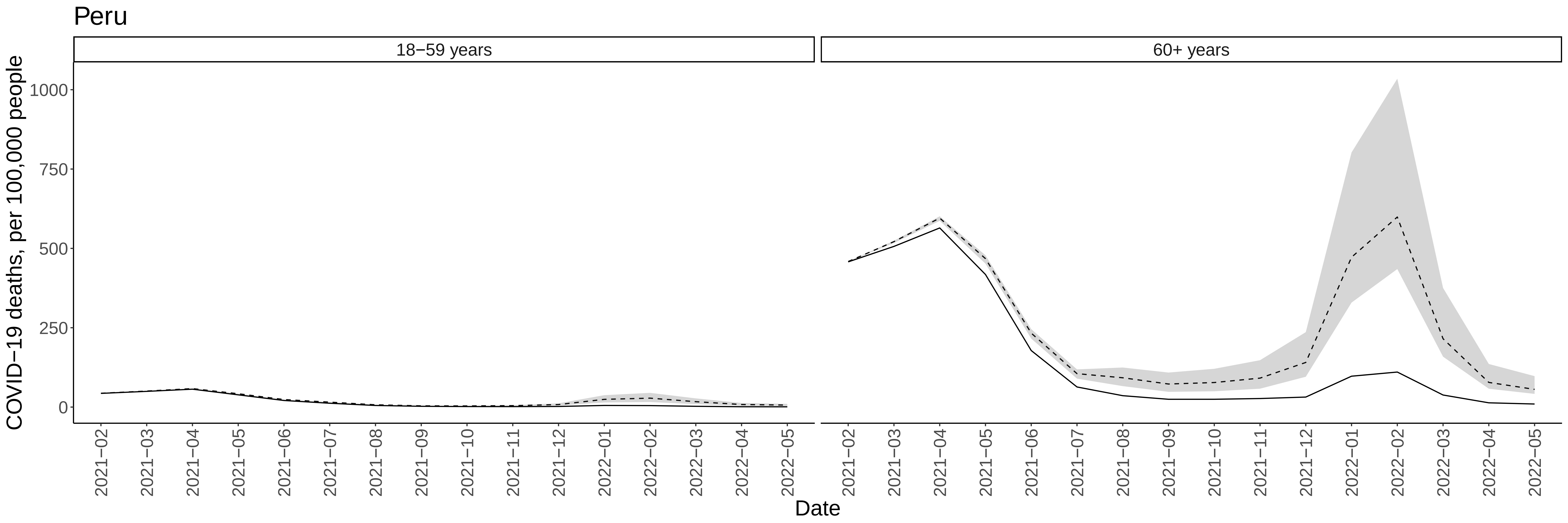

Correction for country−level estimated underreporting of COVID−19 mortality (Msemburi et al, 2023)

Belize

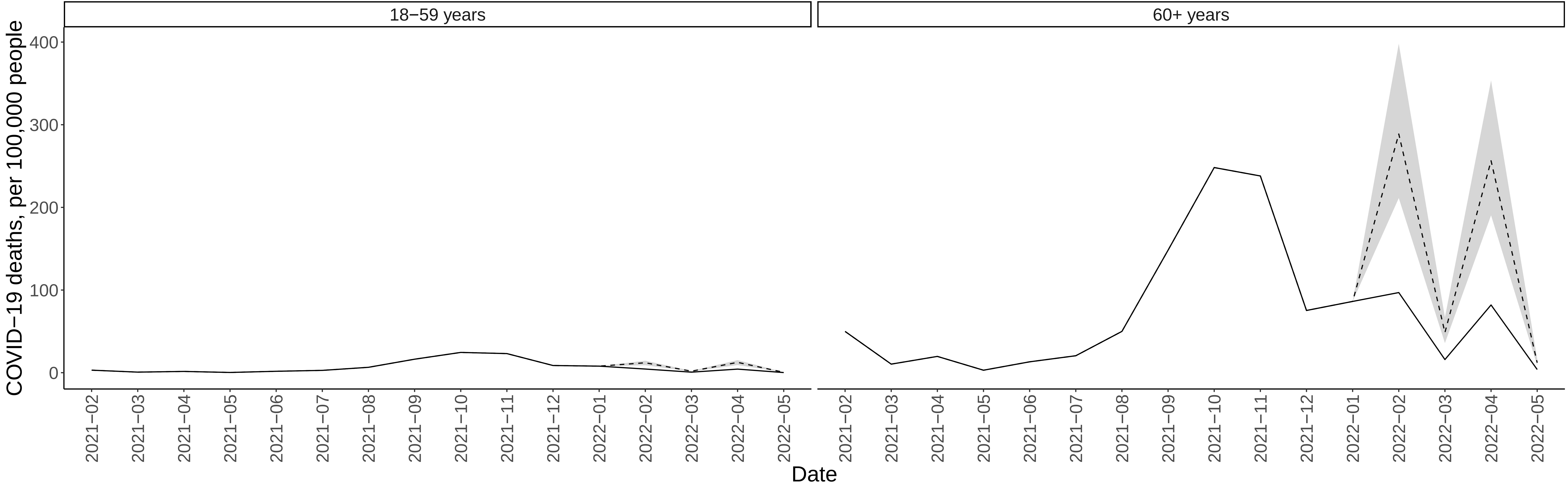


Correction for country−level estimated underreporting of COVID−19 mortality (Msemburi et al, 2023)

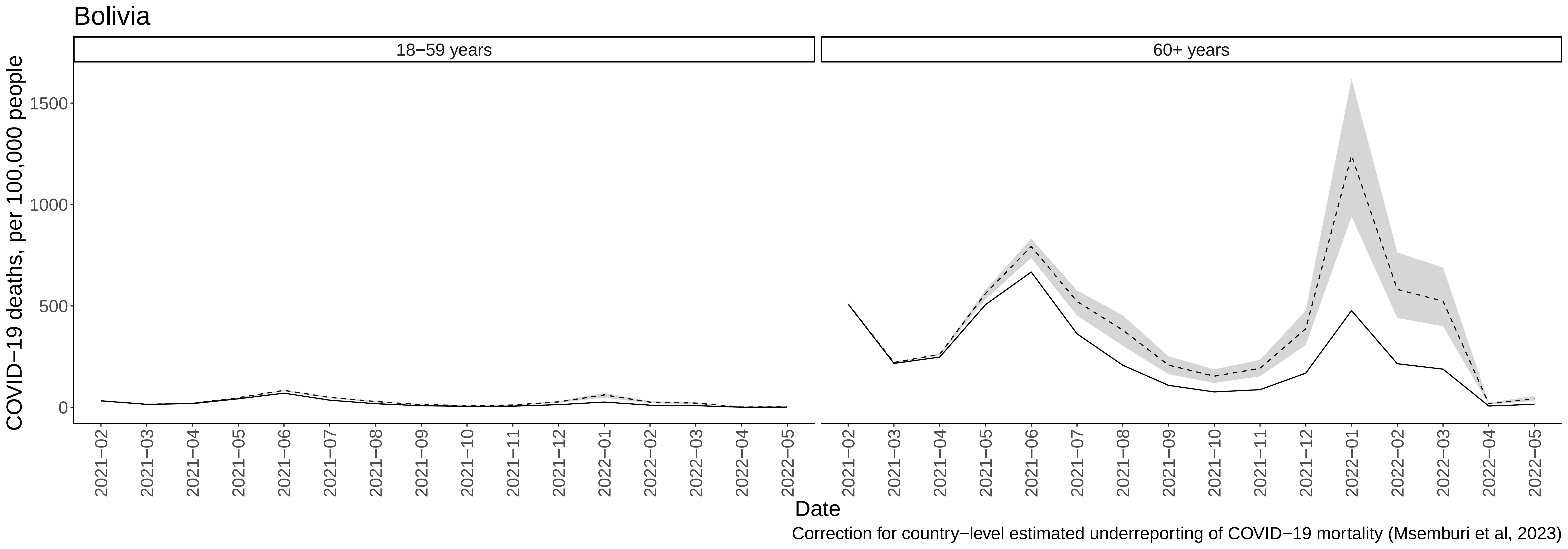

Bolivia
18−59 years
60+ years
COVID−19 deaths, per 100,000 people
0
500
1000
1500
2021−02
2021−03
2021−04
2021−05
2021−06
2021−07
2021−08
2021−09
2021−10
2021−11
2021−12
2022−01
2022−02
2022−03
2022−04
2022−05
Date
Correction for country−level estimated underreporting of COVID−19 mortality (Msemburi et al, 2023)

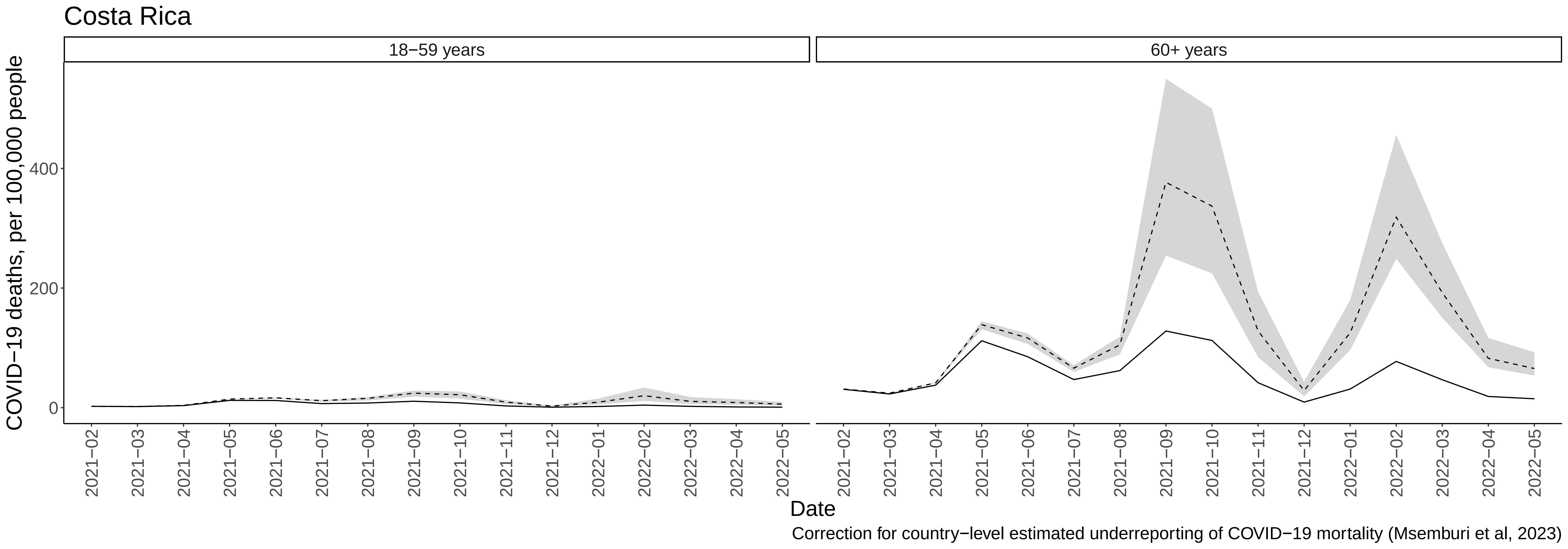
Costa Rica
18−59 years
60+ years
COVID−19 deaths, per 100,000 people
0
200
400
2021−02
2021−03
2021−04
2021−05
2021−06
2021−07
2021−08
2021−09
2021−10
2021−11
2021−12
2022−01
2022−02
2022−03
2022−04
2022−05
Date
Correction for country−level estimated underreporting of COVID−19 mortality (Msemburi et al, 2023)

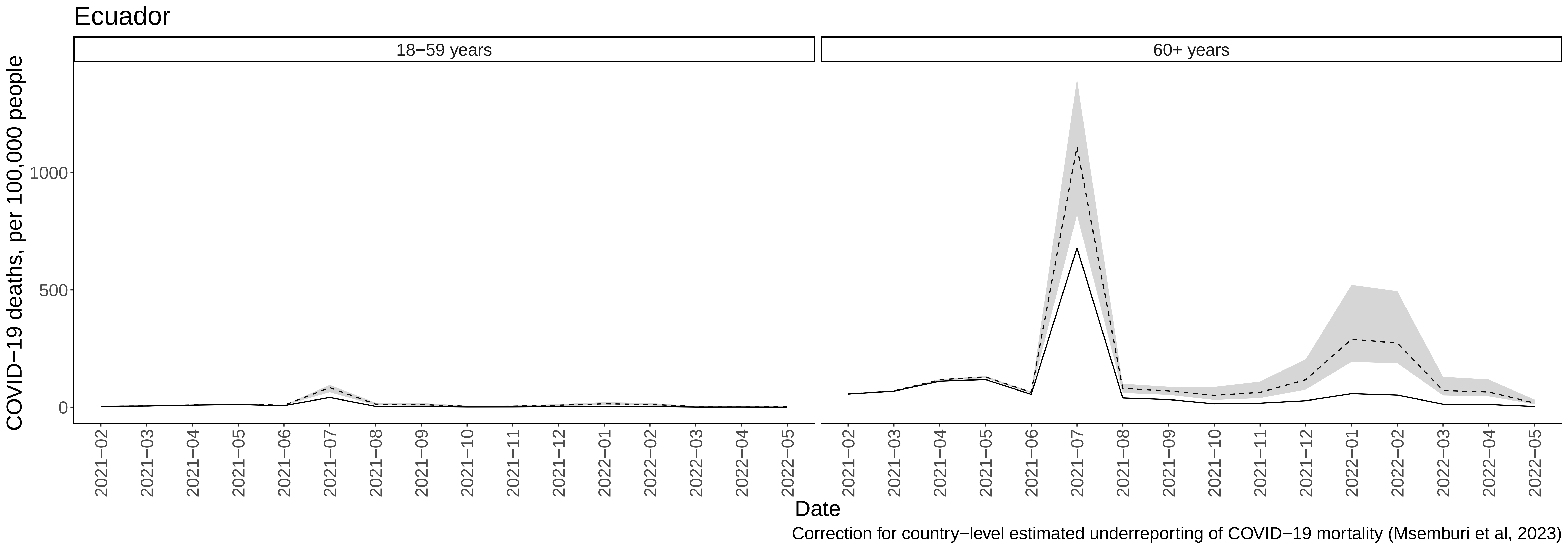

Ecuador
18−59 years
60+ years
COVID−19 deaths, per 100,000 people
0
500
1000
2021−02
2021−03
2021−04
2021−05
2021−06
2021−07
2021−08
2021−09
2021−10
2021−11
2021−12
2022−01
2022−02
2022−03
2022−04
2022−05
Date
Correction for country−level estimated underreporting of COVID−19 mortality (Msemburi et al, 2023)

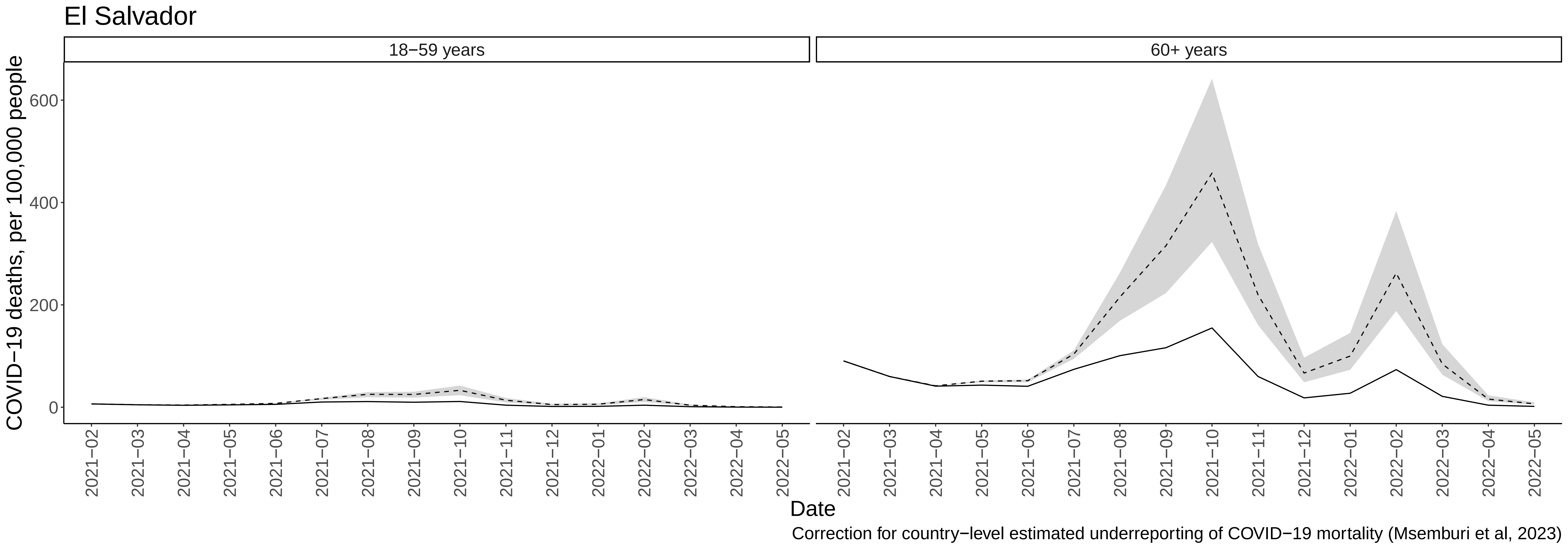
El Salvador
18−59 years
60+ years
COVID−19 deaths, per 100,000 people
0
200
400
600
2021−02
2021−03
2021−04
2021−05
2021−06
2021−07
2021−08
2021−09
2021−10
2021−11
2021−12
2022−01
2022−02
2022−03
2022−04
2022−05
Date
Correction for country−level estimated underreporting of COVID−19 mortality (Msemburi et al, 2023)

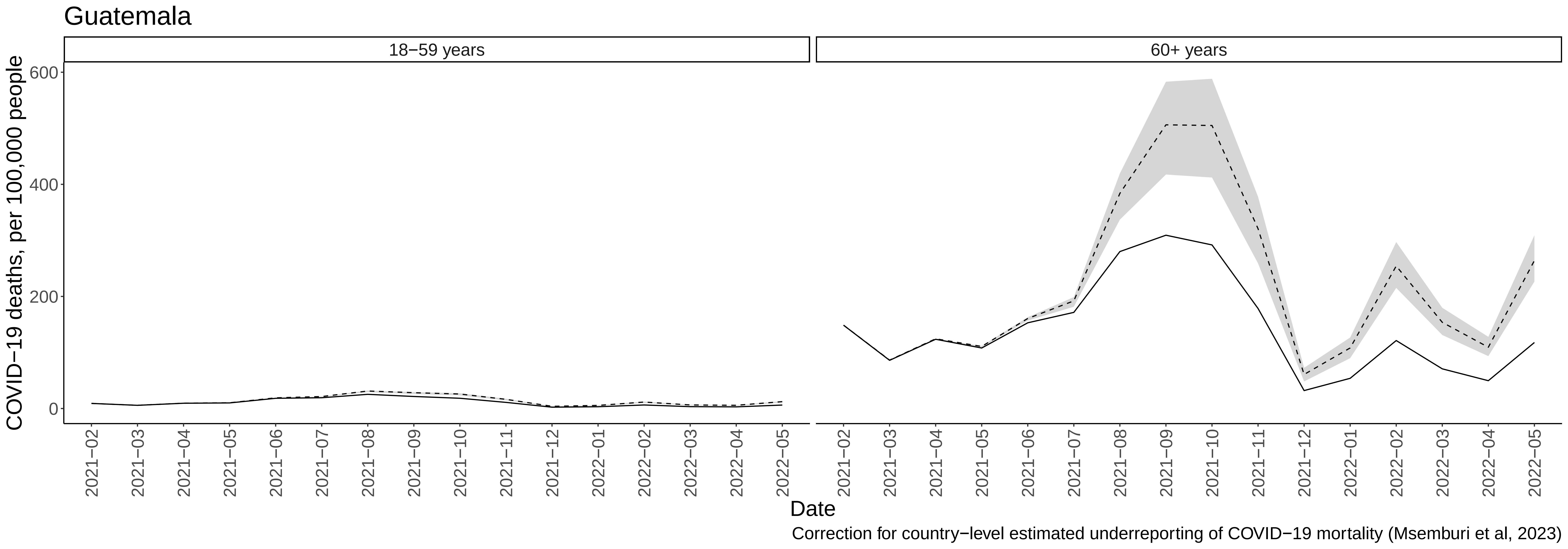

Guatemala
18−59 years
60+ years
COVID−19 deaths, per 100,000 people
600
400
200
0
2021−02
2021−03
2021−04
2021−05
2021−06
2021−07
2021−08
2021−09
2021−10
2021−11
2021−12
2022−01
2022−02
2022−03
2022−04
2022−05
Date
Correction for country−level estimated underreporting of COVID−19 mortality (Msemburi et al, 2023)

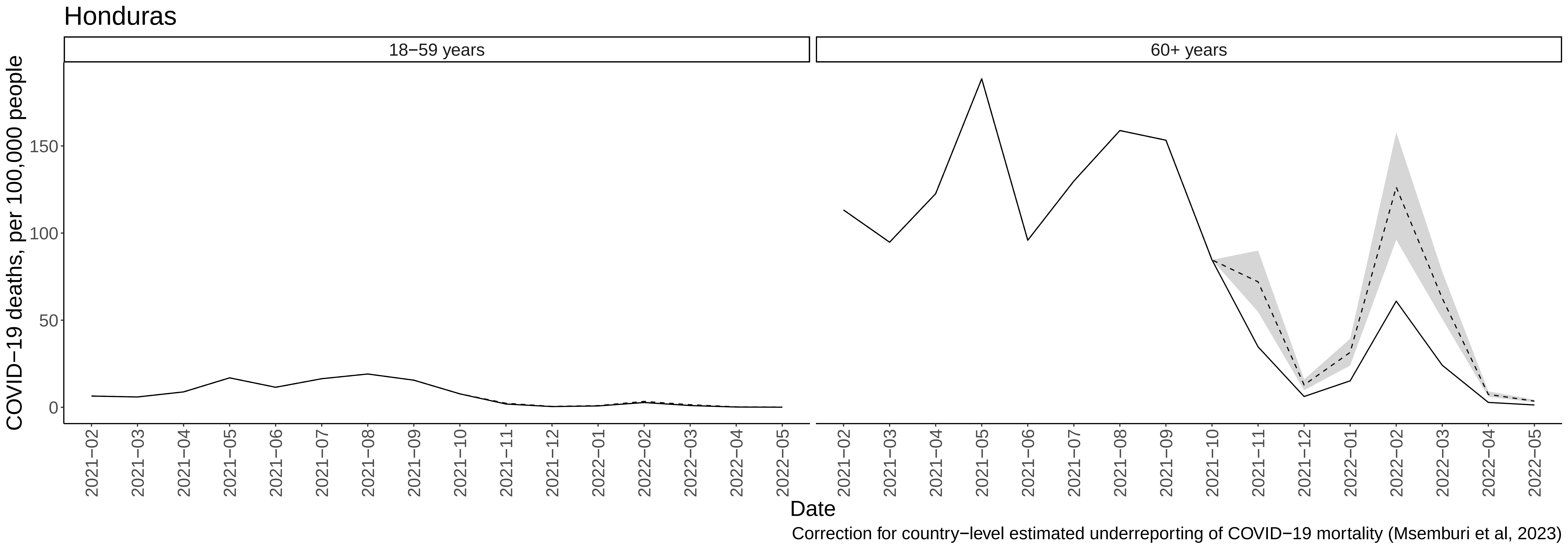

Honduras
18−59 years
60+ years
COVID−19 deaths, per 100,000 people
0
50
100
150
2021−02
2021−03
2021−04
2021−05
2021−06
2021−07
2021−08
2021−09
2021−10
2021−11
2021−12
2022−01
2022−02
2022−03
2022−04
2022−05
Date
Correction for country−level estimated underreporting of COVID−19 mortality (Msemburi et al, 2023)

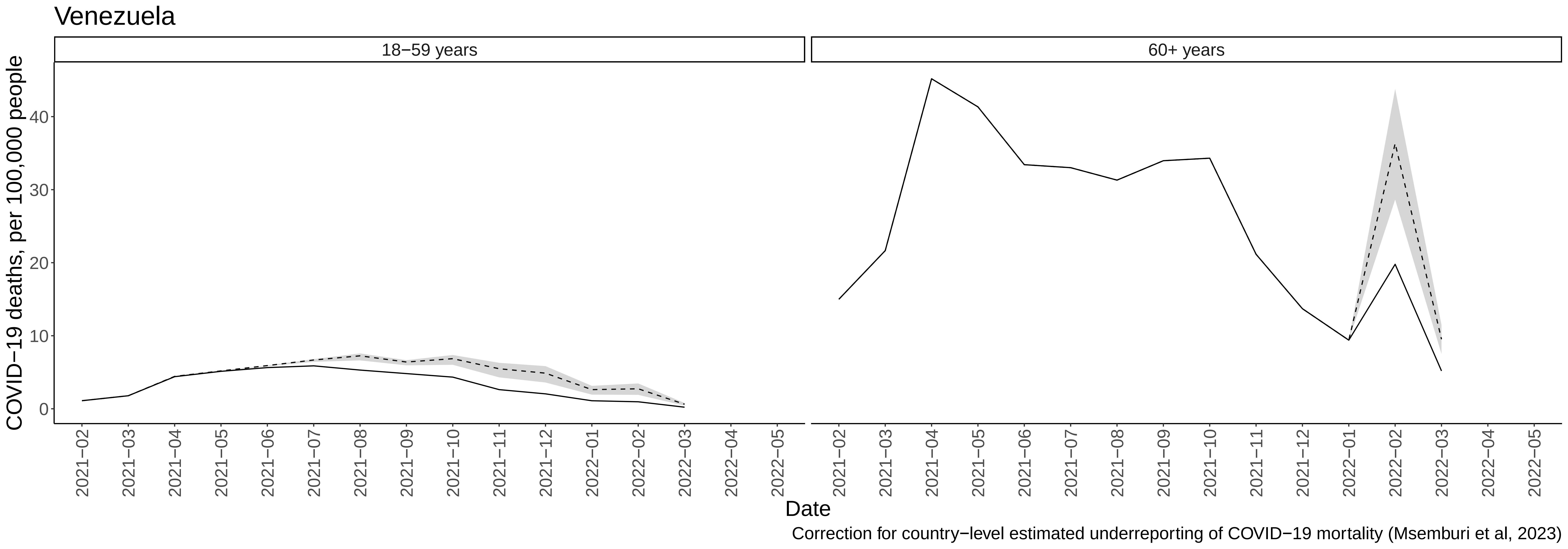
Venezuela
18−59 years
60+ years
COVID−19 deaths, per 100,000 people
0
10
20
30
40
2021−02
2021−03
2021−04
2021−05
2021−06
2021−07
2021−08
2021−09
2021−10
2021−11
2021−12
2022−01
2022−02
2022−03
2022−04
2022−05
Date
Correction for country−level estimated underreporting of COVID−19 mortality (Msemburi et al, 2023)

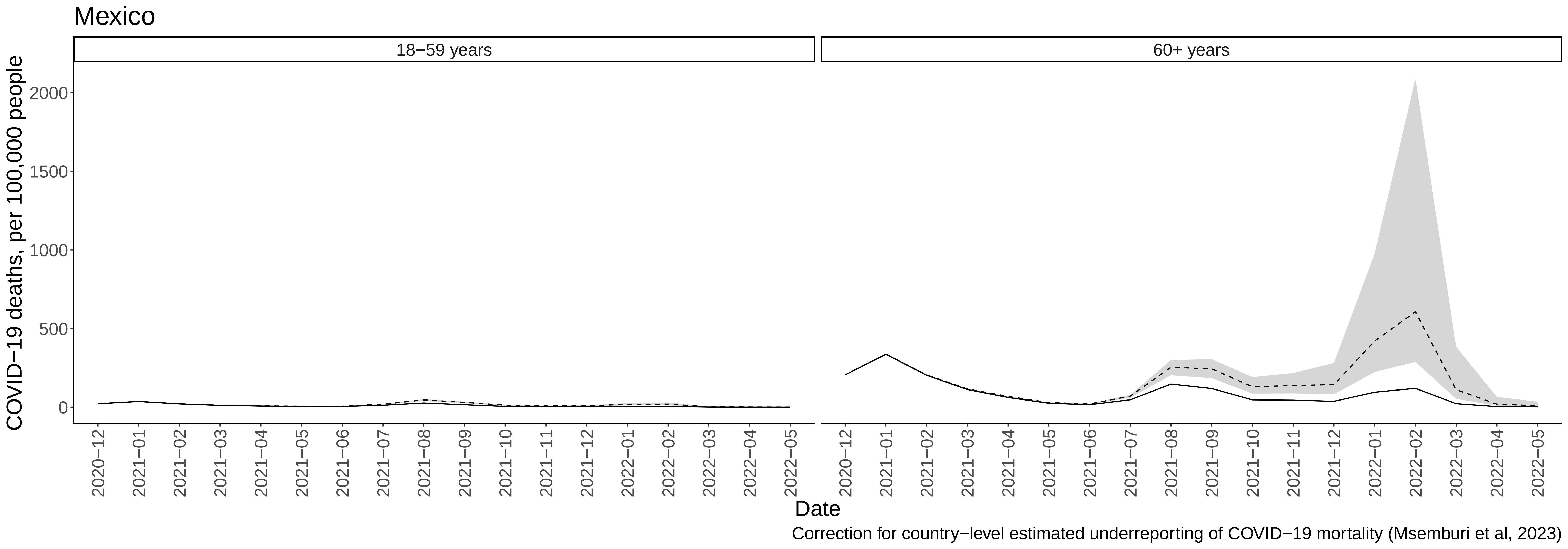

Mexico
18−59 years
60+ years
COVID−19 deaths, per 100,000 people
0
500
1000
1500
2000
2020−12
2021−01
2021−02
2021−03
2021−04
2021−05
2021−06
2021−07
2021−08
2021−09
2021−10
2021−11
2021−12
2022−01
2022−02
2022−03
2022−04
2022−05
Date
Correction for country−level estimated underreporting of COVID−19 mortality (Msemburi et al, 2023)
