## Supplemental Figure 3 for "Deaths averted by COVID-19 vaccination in select Latin American and Caribbean Countries: a modelling study"

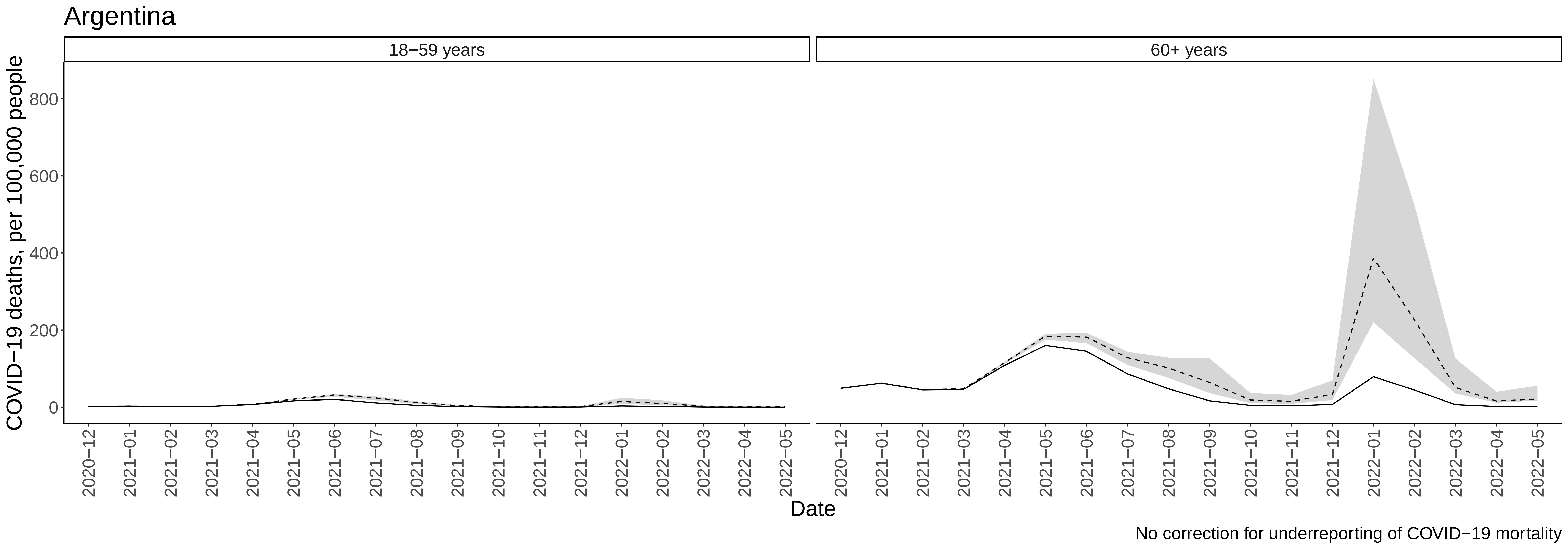

Argentina
18−59 years
60+ years
COVID−19 deaths, per 100,000 people
0
200
400
600
800
2020−12
2021−01
2021−02
2021−03
2021−04
2021−05
2021−06
2021−07
2021−08
2021−09
2021−10
2021−11
2021−12
2022−01
2022−02
2022−03
2022−04
2022−05
Date
No correction for underreporting of COVID−19 mortality

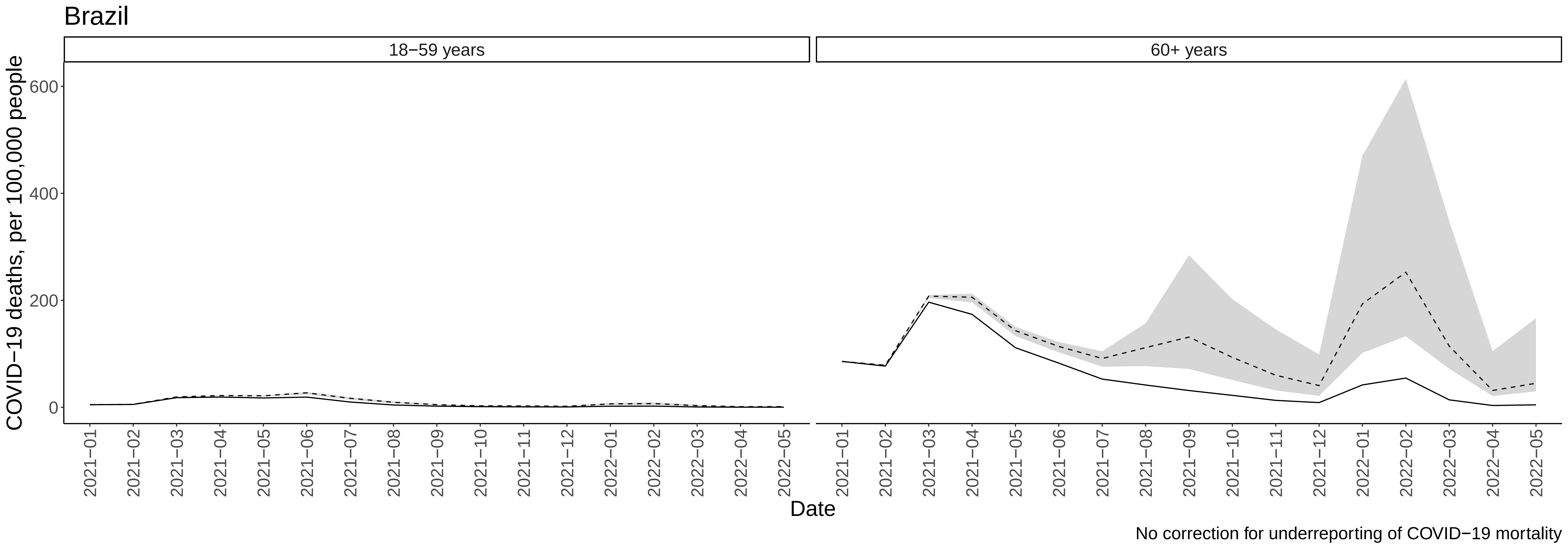
Brazil
18−59 years
60+ years
COVID−19 deaths, per 100,000 people
0
200
400
600
2021−01
2021−02
2021−03
2021−04
2021−05
2021−06
2021−07
2021−08
2021−09
2021−10
2021−11
2021−12
2022−01
2022−02
2022−03
2022−04
2022−05
Date
No correction for underreporting of COVID−19 mortality

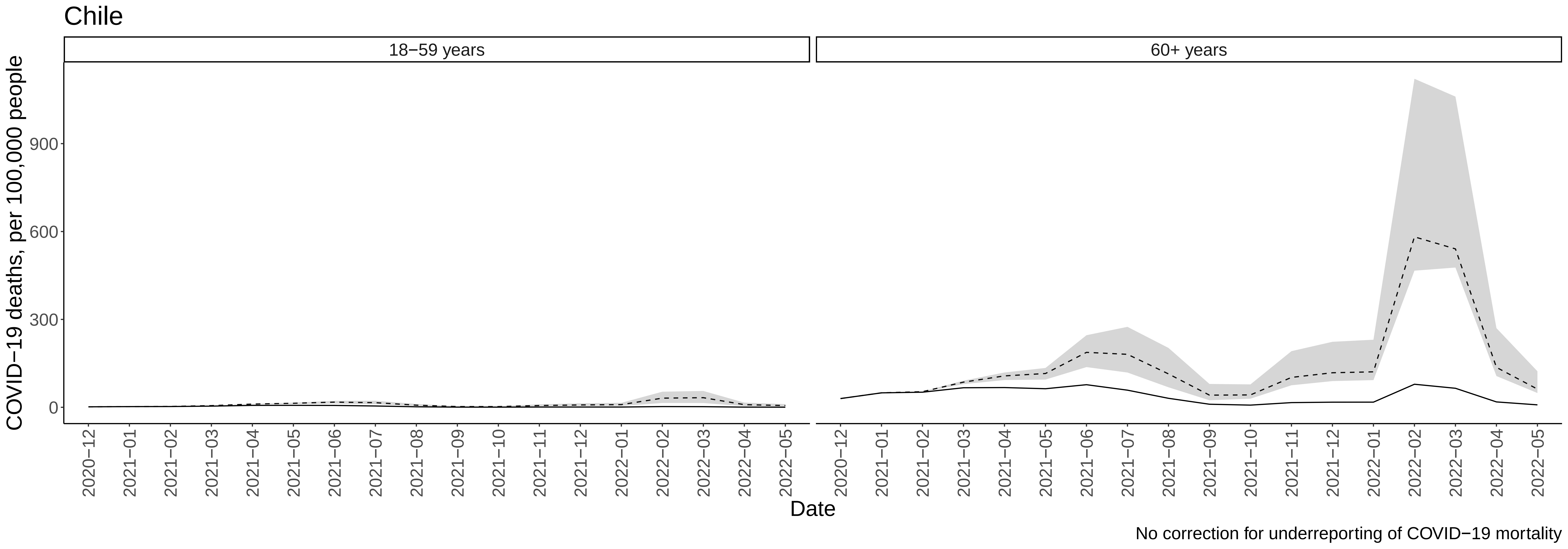
Chile
18−59 years
60+ years
COVID−19 deaths, per 100,000 people
0
300
600
900
2020−12
2021−01
2021−02
2021−03
2021−04
2021−05
2021−06
2021−07
2021−08
2021−09
2021−10
2021−11
2021−12
2022−01
2022−02
2022−03
2022−04
2022−05
Date
No correction for underreporting of COVID−19 mortality

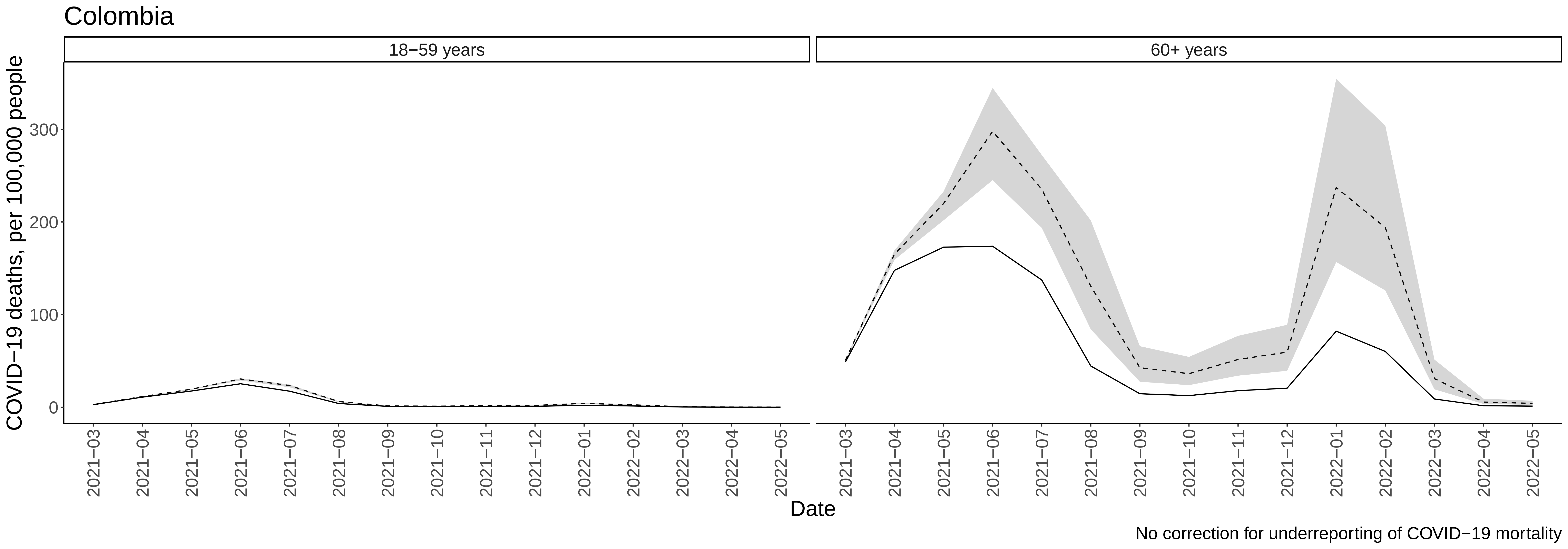

Colombia
18−59 years
60+ years
COVID−19 deaths, per 100,000 people
0
100
200
300
2021−03
2021−04
2021−05
2021−06
2021−07
2021−08
2021−09
2021−10
2021−11
2021−12
2022−01
2022−02
2022−03
2022−04
2022−05
Date
No correction for underreporting of COVID−19 mortality

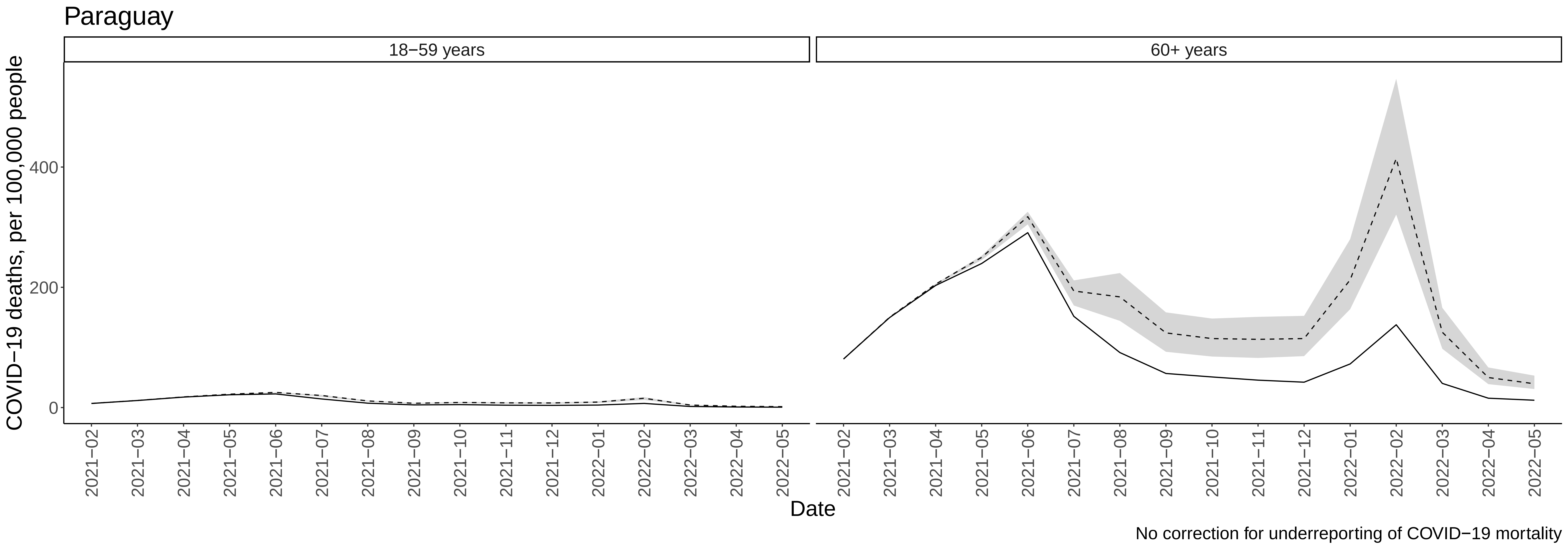
Paraguay
18−59 years
60+ years
COVID−19 deaths, per 100,000 people
0
200
400
2021−02
2021−03
2021−04
2021−05
2021−06
2021−07
2021−08
2021−09
2021−10
2021−11
2021−12
2022−01
2022−02
2022−03
2022−04
2022−05
Date
No correction for underreporting of COVID−19 mortality

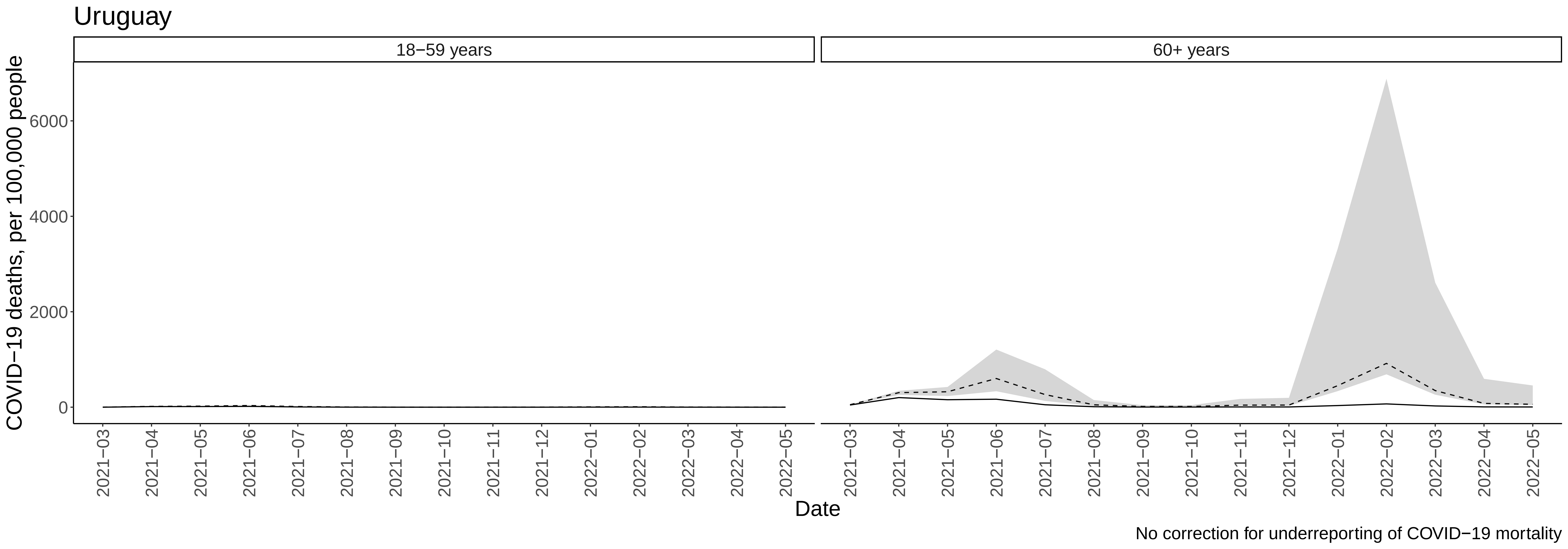
Uruguay
18−59 years
60+ years
COVID−19 deaths, per 100,000 people
0
2000
4000
6000
2021−03
2021−04
2021−05
2021−06
2021−07
2021−08
2021−09
2021−10
2021−11
2021−12
2022−01
2022−02
2022−03
2022−04
2022−05
Date
No correction for underreporting of COVID−19 mortality

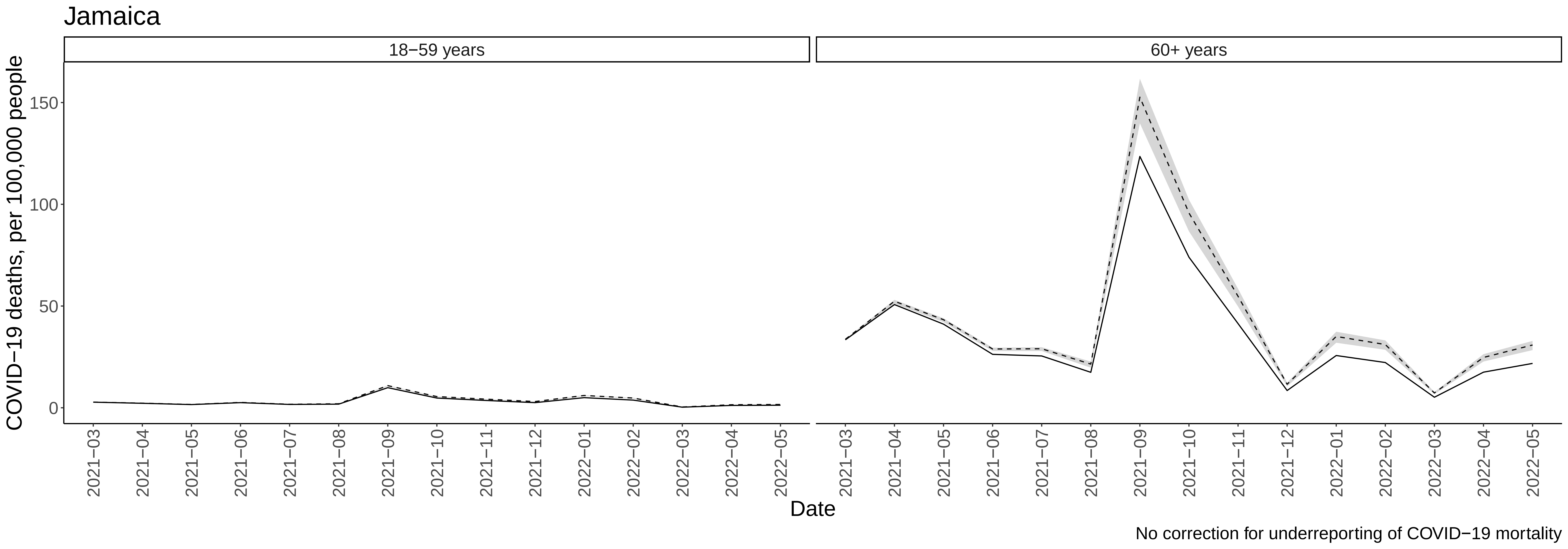
Jamaica
18−59 years
60+ years
COVID−19 deaths, per 100,000 people
0
50
100
150
2021−03
2021−04
2021−05
2021−06
2021−07
2021−08
2021−09
2021−10
2021−11
2021−12
2022−01
2022−02
2022−03
2022−04
2022−05
Date
No correction for underreporting of COVID−19 mortality

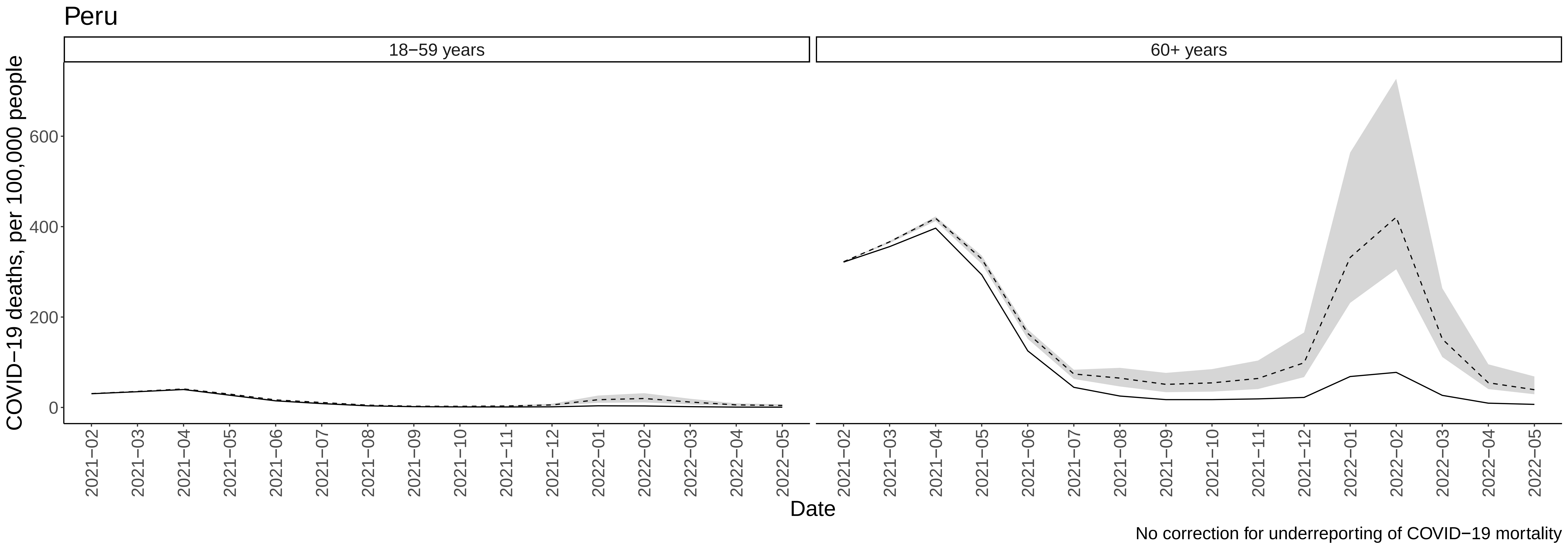
Peru
18−59 years
60+ years
COVID−19 deaths, per 100,000 people
0
200
400
600
2021−02
2021−03
2021−04
2021−05
2021−06
2021−07
2021−08
2021−09
2021−10
2021−11
2021−12
2022−01
2022−02
2022−03
2022−04
2022−05
Date
No correction for underreporting of COVID−19 mortality

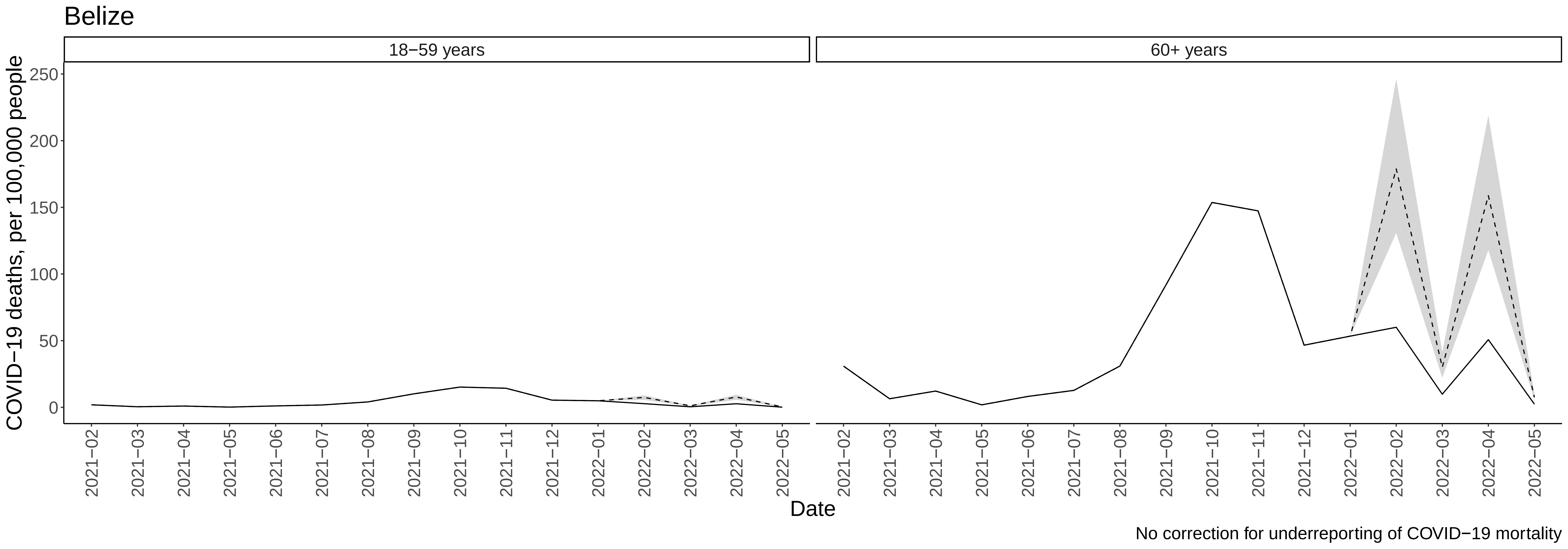
Belize
18−59 years
60+ years
COVID−19 deaths, per 100,000 people
250
200
150
100
50
0
2021−02
2021−03
2021−04
2021−05
2021−06
2021−07
2021−08
2021−09
2021−10
2021−11
2021−12
2022−01
2022−02
2022−03
2022−04
2022−05
Date
No correction for underreporting of COVID−19 mortality

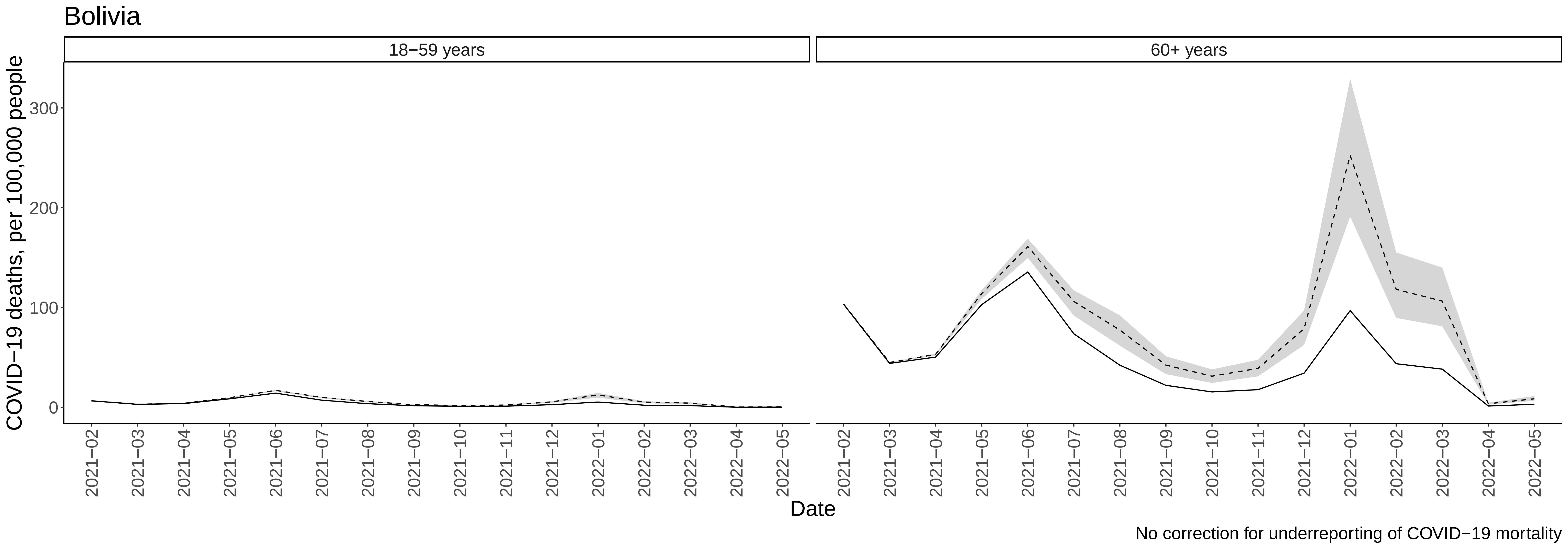

Bolivia
18−59 years
60+ years
COVID−19 deaths, per 100,000 people
0
100
200
300
2021−02
2021−03
2021−04
2021−05
2021−06
2021−07
2021−08
2021−09
2021−10
2021−11
2021−12
2022−01
2022−02
2022−03
2022−04
2022−05
Date
No correction for underreporting of COVID−19 mortality

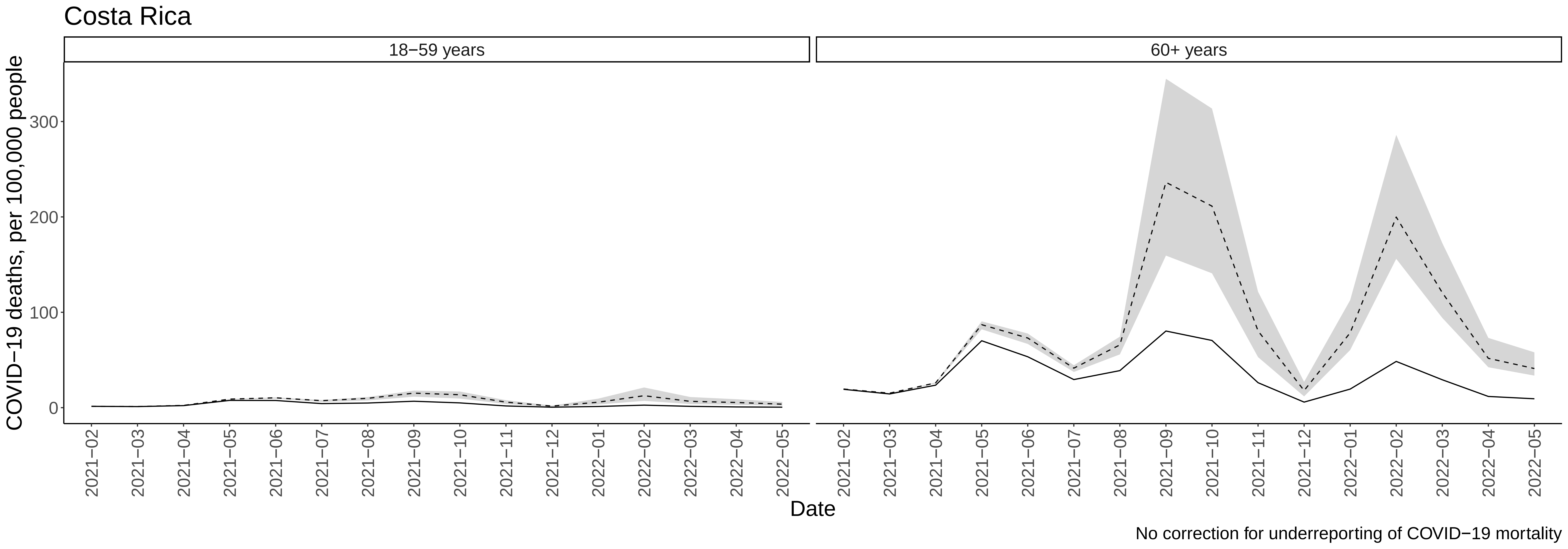

Costa Rica
18−59 years
60+ years
COVID−19 deaths, per 100,000 people
0
100
200
300
2021−02
2021−03
2021−04
2021−05
2021−06
2021−07
2021−08
2021−09
2021−10
2021−11
2021−12
2022−01
2022−02
2022−03
2022−04
2022−05
Date
No correction for underreporting of COVID−19 mortality

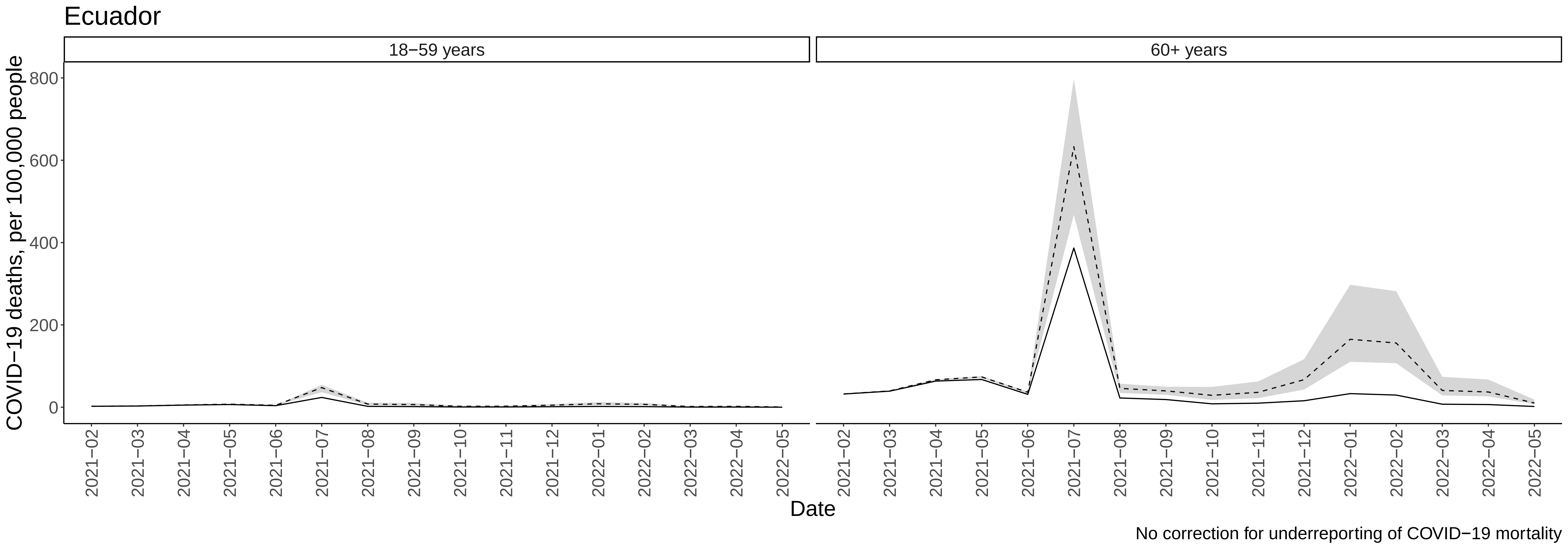

Ecuador
18−59 years
60+ years
COVID−19 deaths, per 100,000 people
800
600
400
200
0
2021−02
2021−03
2021−04
2021−05
2021−06
2021−07
2021−08
2021−09
2021−10
2021−11
2021−12
2022−01
2022−02
2022−03
2022−04
2022−05
Date
No correction for underreporting of COVID−19 mortality

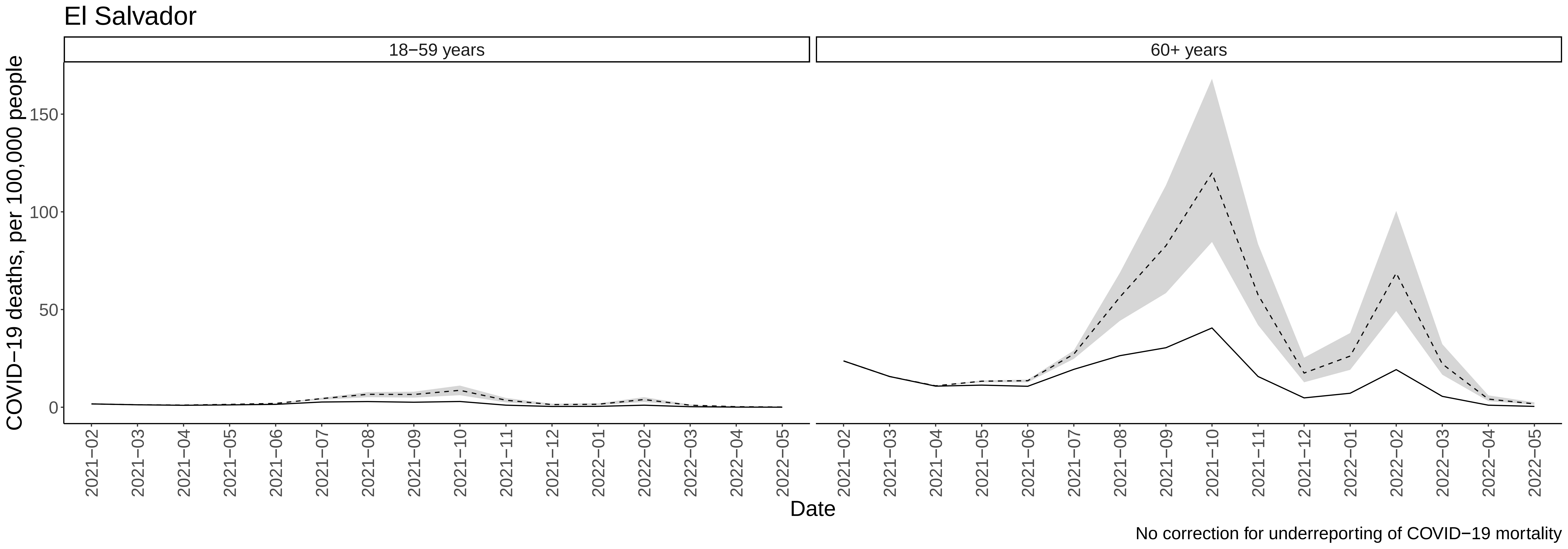

El Salvador
18−59 years
60+ years
COVID−19 deaths, per 100,000 people
0
50
100
150
2021−02
2021−03
2021−04
2021−05
2021−06
2021−07
2021−08
2021−09
2021−10
2021−11
2021−12
2022−01
2022−02
2022−03
2022−04
2022−05
Date
No correction for underreporting of COVID−19 mortality

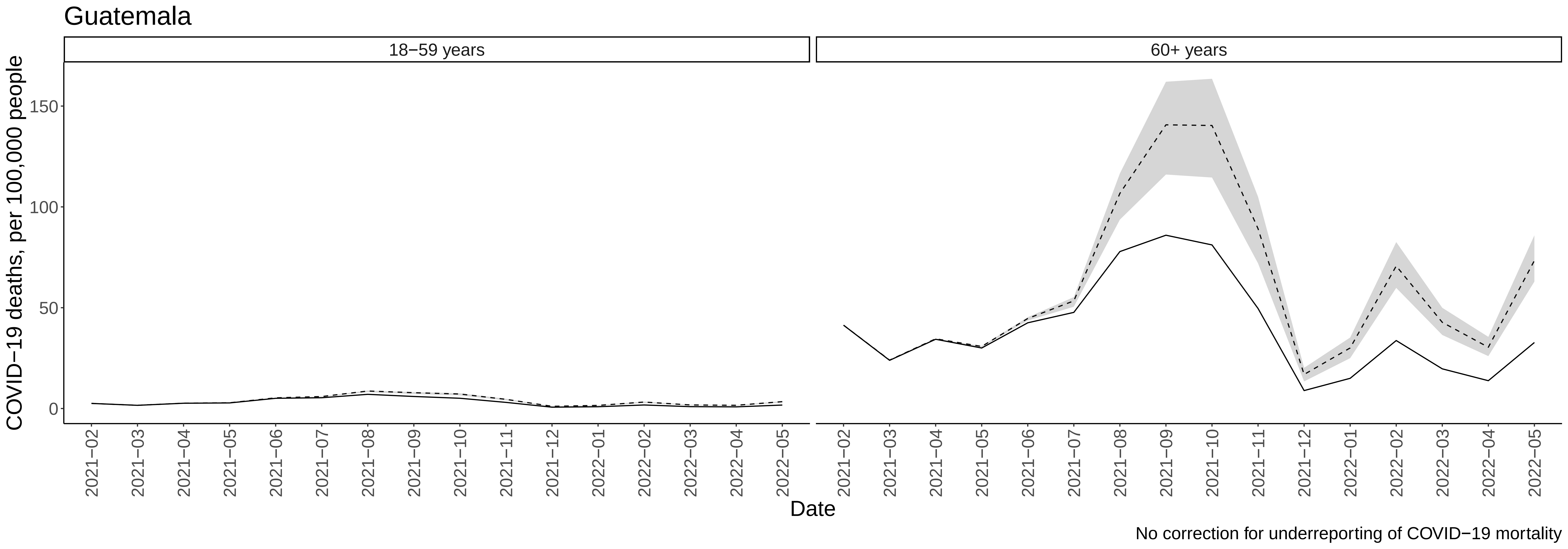

Guatemala
18−59 years
60+ years
COVID−19 deaths, per 100,000 people
0
50
100
150
2021−02
2021−03
2021−04
2021−05
2021−06
2021−07
2021−08
2021−09
2021−10
2021−11
2021−12
2022−01
2022−02
2022−03
2022−04
2022−05
Date
No correction for underreporting of COVID−19 mortality

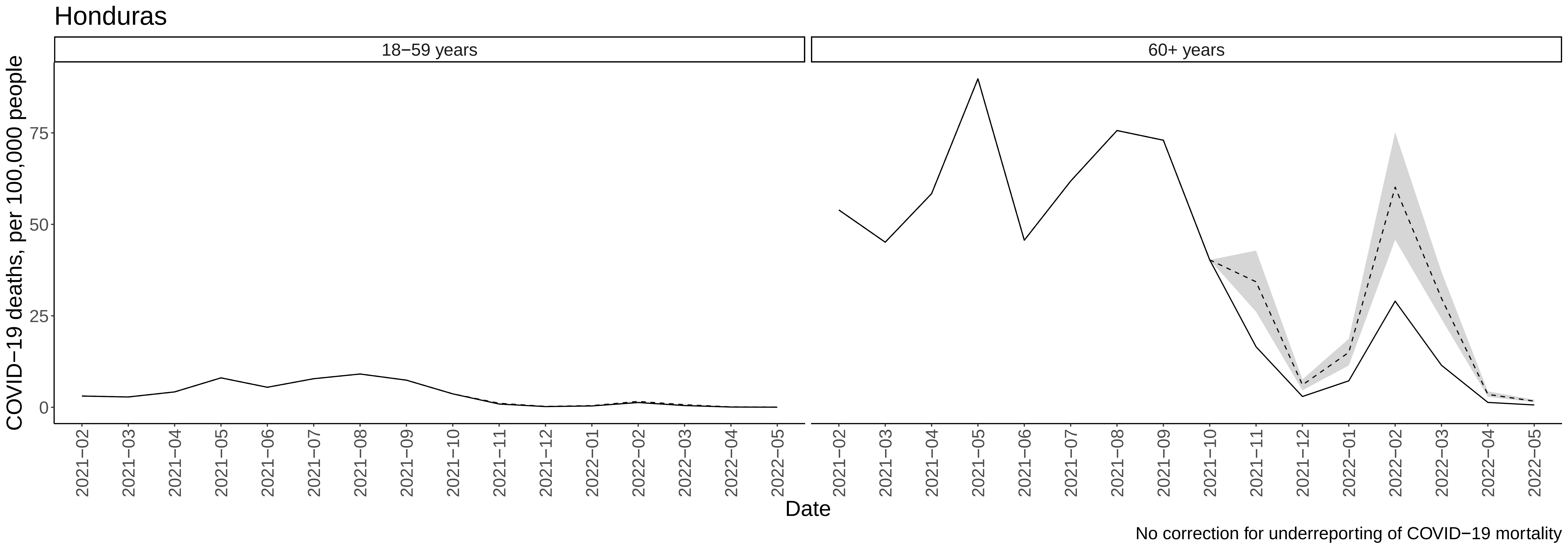

Honduras
18−59 years
60+ years
COVID−19 deaths, per 100,000 people
0
25
50
75
2021−02
2021−03
2021−04
2021−05
2021−06
2021−07
2021−08
2021−09
2021−10
2021−11
2021−12
2022−01
2022−02
2022−03
2022−04
2022−05
Date
No correction for underreporting of COVID−19 mortality

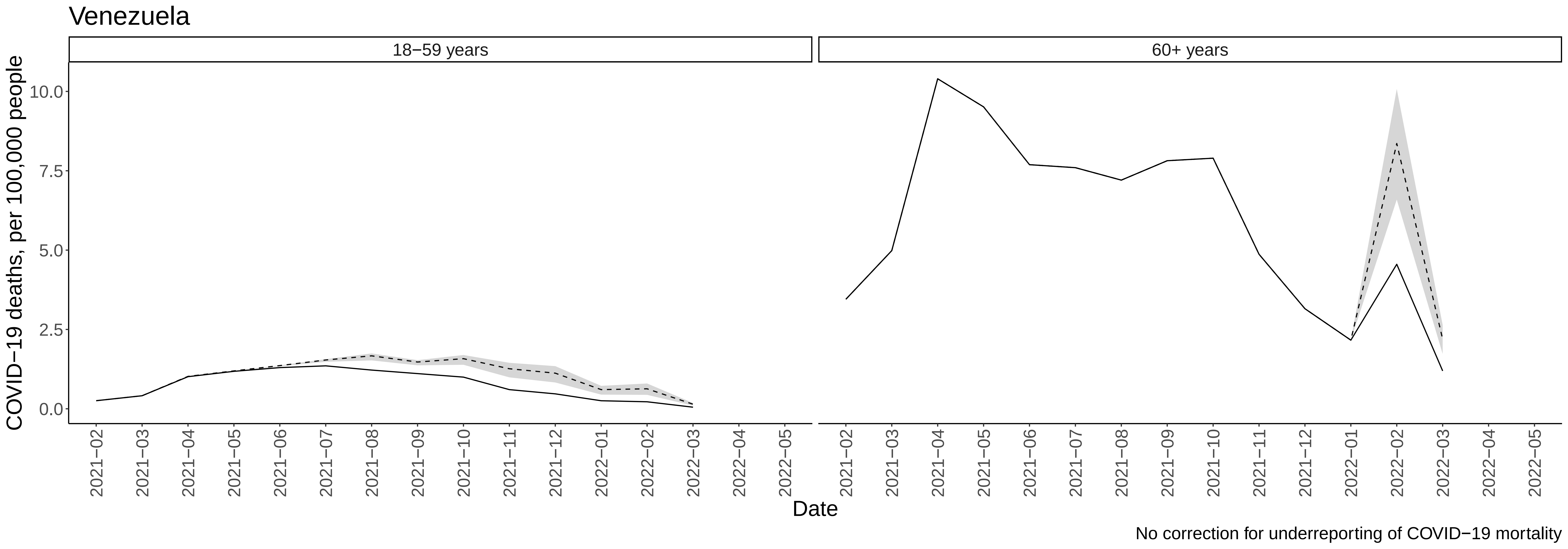
Venezuela
18−59 years
60+ years
COVID−19 deaths, per 100,000 people
0.0
2.5
5.0
7.5
10.0
2021−02
2021−03
2021−04
2021−05
2021−06
2021−07
2021−08
2021−09
2021−10
2021−11
2021−12
2022−01
2022−02
2022−03
2022−04
2022−05
Date
No correction for underreporting of COVID−19 mortality

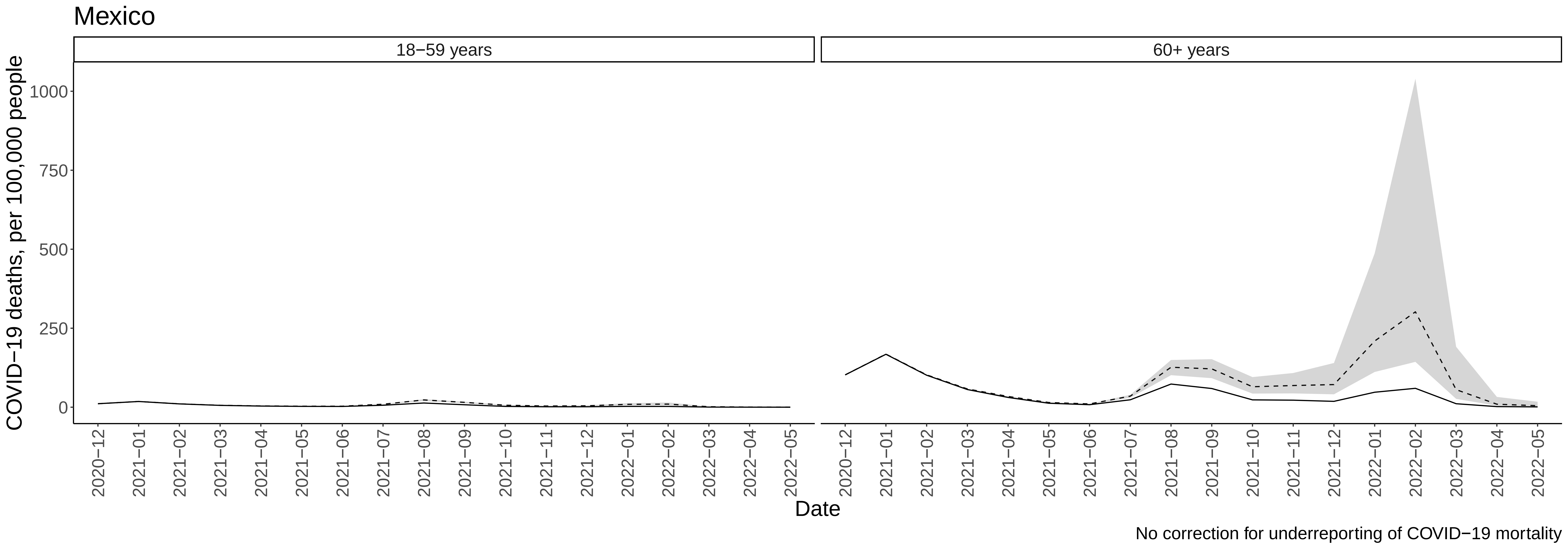

Mexico
18−59 years
60+ years
COVID−19 deaths, per 100,000 people
1000
750
500
250
0
2020−12
2021−01
2021−02
2021−03
2021−04
2021−05
2021−06
2021−07
2021−08
2021−09
2021−10
2021−11
2021−12
2022−01
2022−02
2022−03
2022−04
2022−05
Date
No correction for underreporting of COVID−19 mortality
