## Supplementary Tables for "Deaths averted by COVID-19 vaccination in select Latin American and Caribbean Countries: a modelling study"

|  | With adjustment for under-reporting (country-specific) | | | | No adjustment for under-reporting | | | | |
| --- | --- | --- | --- | --- | --- | --- | --- | --- | --- |
| Country | Observed deaths, per 100,000 population | Deaths averted, medium vaccine effectiveness, per 100,000 population | Deaths averted, low vaccine effectiveness, per 100,000 population | Deaths averted, high vaccine effectiveness, per 100,000 population | | Observed deaths, per 100,000 population | Deaths averted, medium vaccine effectiveness, per 100,000 population | Deaths averted, low vaccine effectiveness, per 100,000 population | Deaths averted, high vaccine effectiveness, per 100,000 population |
| Argentina | 264.9 | 237.98 | 118.68 | 508.77 | | 264.9 | 108.56 | 50.91 | 298.39 |
| Brazil | 309.0 | 250.15 | 123.83 | 589.95 | | 279.9 | 98.47 | 43.02 | 251.02 |
| Chile | 216.5 | 588.29 | 395.26 | 1133.09 | | 202.8 | 354.42 | 235.18 | 866.29 |
| Colombia | 301.9 | 207.48 | 110.58 | 330.17 | | 238 | 74.78 | 28.17 | 150.87 |
| Paraguay | 424.4 | 220.53 | 136.41 | 311.47 | | 351.2 | 104.58 | 56.84 | 188.70 |
| Uruguay | 252.1 | 795.24 | 485.55 | 4507.62 | | 252.1 | 462.61 | 259.98 | 2133.38 |
| Jamaica | 250.1 | 47.27 | 33.09 | 61.45 | | 120.6 | 14.18 | 4.73 | 23.63 |
| Peru | 656.9 | 372.24 | 226.21 | 651.75 | | 461.6 | 144.30 | 84.50 | 351.01 |
| Belize | 230.8 | 75.82 | 37.91 | 113.74 | | 142.9 | 37.91 | 0.00 | 37.91 |
| Bolivia | 757.8 | 428.42 | 262.84 | 595.33 | | 154 | 49.94 | 27.60 | 91.99 |
| Costa Rica | 242.4 | 345.76 | 203.84 | 549.60 | | 152 | 116.11 | 61.93 | 283.83 |
| Ecuador | 298.5 | 294.74 | 147.37 | 511.70 | | 170.1 | 81.05 | 42.57 | 207.96 |
| El Salvador | 221.2 | 280.67 | 165.64 | 427.91 | | 57.9 | 36.81 | 20.71 | 87.42 |
| Guatemala | 421.1 | 183.54 | 117.05 | 241.60 | | 117 | 30.90 | 16.86 | 51.50 |
| Honduras | 236.8 | 20.02 | 12.32 | 27.72 | | 112.8 | 4.62 | 1.54 | 9.24 |
| Venezuela | 105.9 | 18.24 | 10.73 | 24.14 | | 24.4 | 2.68 | 2.15 | 5.36 |
| Mexico | 433.0 | 331.64 | 149.50 | 837.39 | | 215.5 | 63.23 | 28.79 | 175.36 |

**Supplementary Table 1.** Sensitivity analysis with observed and averted COVID-19 deaths via vaccination, by country, considering varying estimates of vaccine effectiveness, and adjustment for mortality under-reporting.
